# Dietary sulfur amino acid restriction in humans with overweight and obesity: a translational randomized controlled trial

**DOI:** 10.1101/2023.09.09.23295155

**Authors:** Thomas Olsen, Emma Stolt, Bente Øvrebø, Amany Elshorbagy, Elena C. Tore, Sindre Lee-Ødegård, Hannibal Troensegaard, Hanna Johannessen, Beate Doeland, Anna A. D. Vo, Anja F. Dahl, Karianne Svendsen, Magne Thoresen, Helga Refsum, Russell Rising, Kristýna Barvíková, Marleen van Greevenbroek, Viktor Kožich, Kjetil Retterstøl, Kathrine J. Vinknes

**Affiliations:** Department of Nutrition, Institute of Basic Medical Sciences, University of Oslo, Oslo, Norway; Department of Food Safety, Norwegian Institute of Public Health, Oslo, Norway; Department of Physiology, Faculty of Medicine, University of Alexandria, Alexandria, Egypt; Department of Pharmacology, University of Oxford, UK; Department of Internal Medicine and CARIM School of Cardiovascular Diseases, Maastricht University, Maastricht, The Netherlands; Department of Clinical Medicine, Faculty of Medicine, University of Oslo; Department of Paedriatic Surgery, Oslo University Hospital, Rikshospitalet, Oslo, Norway; The Lipid Clinic, Department of Endocrinology, Morbid Obesity and Preventive Medicine, Oslo University Hospital, Oslo, Norway; Department of Biostatistics, Institute of Basic Medical Sciences, University of Oslo, Oslo, Norway; D&S Consulting Services, Inc., New York, USA; Department of Pediatrics and Inherited Metabolic Disorders, First Faculty of Medicine, Charles University and General University Hospital, Prague, Czech Republic

**Author notes:** Shared authorship (Thomas Olsen, Emma Stolt). Correspondence: Thomas Olsen.

## Abstract

Dietary sulfur amino acid restriction (SAAR) improves metabolic health in animals, but in humans, SAAR has not been investigated in translational clinical trials. In this study, we investigated the effect of dietary SAAR on body weight, body composition, resting metabolic rate, gene expression profiles in white adipose tissue (WAT), and an extensive blood biomarker profile in 59 humans with overweight and obesity in a double-blind, randomized controlled trial (ClinicalTrials.gov: NCT04701346). Participants were randomized to a plant-based diet low (∼2 g/d, SAAR) or high (∼5.6 g/d, control group) in sulfur amino acids. The diets were provided in full to the participants. After 8 weeks of intervention, SAAR led to a ∼20 % greater weight loss compared to controls (β (95 % CI) -1.14 (-2.04, -0.25) kg, p = 0.012). Despite greater weight loss, resting metabolic rate remained similar between groups. Furthermore, SAAR decreased serum leptin, and increased ketone bodies compared to controls. In WAT, 20 genes were upregulated whereas 24 genes were downregulated (FDR < 5 %) in the SAAR group compared to controls. Generally applicable gene set enrichment analyses revealed that processes associated with ribosomes were upregulated, whereas processer related to structural components were downregulated. In conclusion, our study shows that SAAR leads to weight loss and metabolic benefits. Further research SAAR is needed to investigate the therapeutic potential for metabolic conditions in humans.

## 1. Introduction

Methionine and cysteine are essential and semi-essential sulfur amino acids (SAA) with a FAO/WHO minimum (recommended) intake of 15 (19) mg/kg/d in adults^1^. High dietary intake and plasma concentrations of SAA are strongly and linearly related to increased risk of metabolic disease and development of obesity^2–9^. In a large population-based cohort study of men and women (n = 5179), the difference in fat mass between lower and upper percentiles of plasma total cysteine (tCys) was 11 kg^3^. Moreover, in two independent Dutch cohorts of men and women aged 40–74 y, plasma methionine was associated with liver fat, whereas tCys was associated with overall obesity^9^. Recent prospective cohort studies showed that plasma tCys was associated with diabetes risk^10^, whereas dietary intake of SAA was associated with increased cardiovascular risk scores, diabetes risk and diabetes-related mortality^6–8^.

Dietary sulfur amino acid restriction (SAAR) has been investigated in experimental animal studies for 30 years, and has been shown to have numerous beneficial effects on metabolic health including reduced body weight, improved body composition and insulin sensitivity, and increased energy expenditure^11–21^. In addition, SAAR is associated with changes in several blood biomarkers including decreased leptin and triglycerides, and increased ketone bodies and adiponectin^12–14,22–24^. These SAAR-induced effects are triggered by metabolic adaptions, including mitochondrial biogenesis and transcriptional remodeling of lipid and glucose metabolism in several tissues, but especially in white adipose tissue (WAT) and liver^17^. Important mediators of these effects include the liver hormones fibroblast growth factor 21 (FGF-21)^25,26^ and insulin-like growth factor 1 (IGF-1)^27,28^.

Despite these robust findings in experimental and observational studies, dietary interventions with SAAR in humans at risk of metabolic disease are scarce^29–31^. Of these, only one long-term, double-blind randomized controlled trial has been published^30^. This 16-week study implementing dietary methionine restriction with a mostly powder-based diet demonstrated increased fat oxidation and decreased liver fat. However, there was a lack of adherence and a high dropout-rate partly due to the synthetic nature of the diet.

To increase palatability and adherence to a SAAR regimen, we developed an approach using plant-based whole foods. Results from two double-blind one-week pilot studies using this approach indicated that some metabolic effects on plasma and urinary SAA, FGF-21, and adipose tissue gene expression were apparent in humans within one week^31,32^. Building on previous data from animals and humans, we performed an 8-week double-blind randomized controlled dietary intervention in individuals with overweight and obesity. Our primary aim was to study the effects of SAAR on body weight. In addition, we assessed the effects of SAAR on several exploratory outcomes, including body mass compartments, energy expenditure, metabolic biomarkers, adipokines and appetite-related hormones, and adipose tissue gene expression profiles.

## 2. Results

Baseline characteristics, effect sizes and standard errors (SE) from intention-to-treat (ITT) analyses are presented in **Tables 1** to **4** and **Figures 1** to **3**. Raw observed means and estimated marginal means (EMMs) derived from the regression models for all variables and all study visits are presented in the online supplement for the ITT population (**Supplemental Tables S3** to **S5**). All per-protocol (PP) analyses excluding drop-outs are presented in the online supplement (**Supplemental Tables S6** to **S8**).

### 2.1. Participant flow and baseline characteristics

A detailed participant flow is presented in **Supplemental Figure S1** and a simplified overview in **Figure 1A**. In total, 61 individuals were randomized to SAAR (n = 32) or control (n = 29). Two participants dropped out pre-baseline resulting in a total of 59 participants with baseline visits who were analyzed ITT (SAAR n = 31; controls n = 28). Five additional drop-outs during the course of the trial led to a total of 54 completers who were analyzed PP (**Supplemental Tables S6** to **S8**).

**Figure 1.**
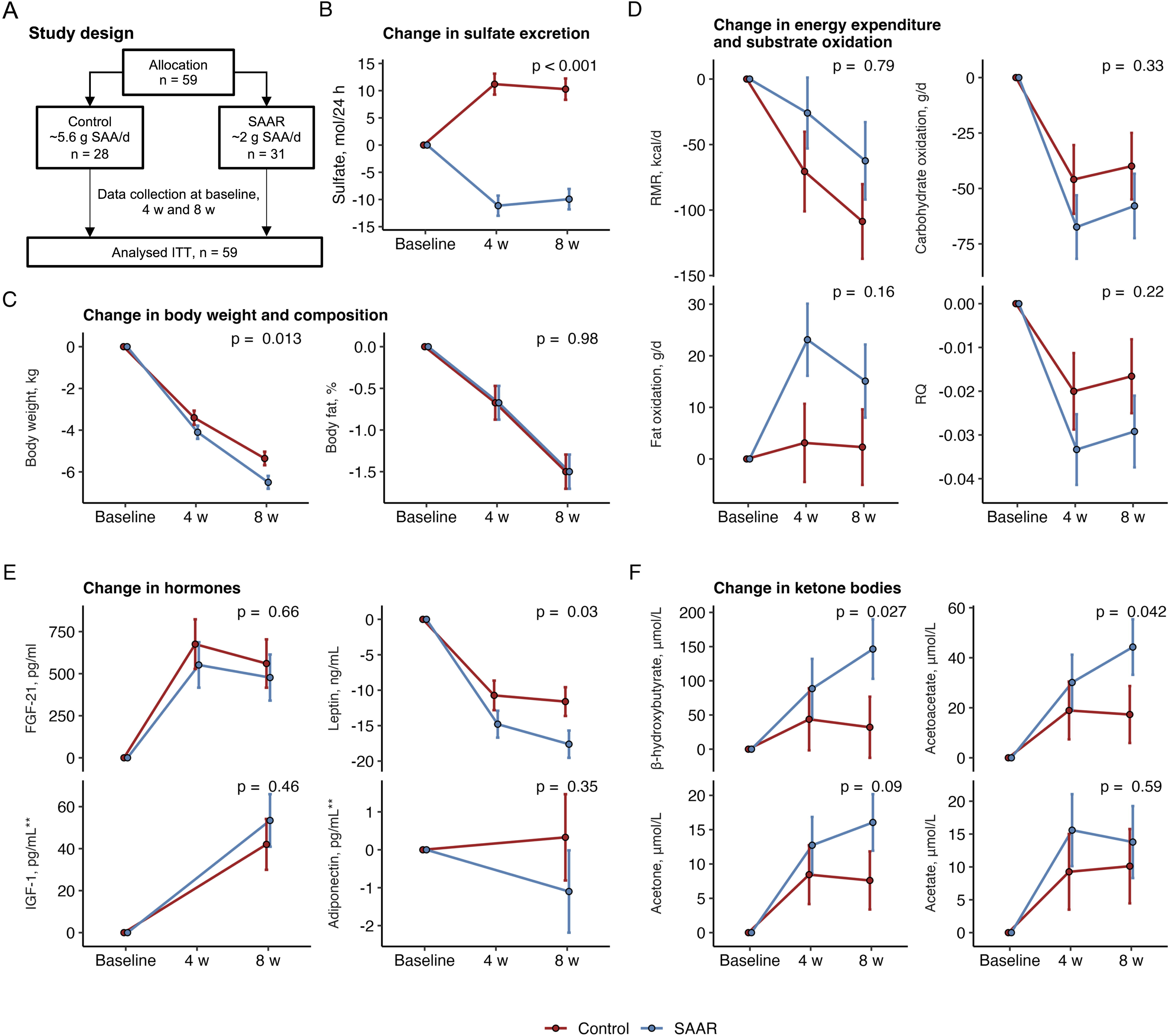
A) Simplified study overview (see full CONSORT diagram in Supplemental Figure S1; Effects of SAAR on B) urinary sulfate excretion; C) Change in body weight and body fat; D) resting metabolic rate (RMR), carbohydrate oxidation, fat oxidation and respiratory quotient (RQ); E) serum levels of fibroblast growth factor-21 (FGF-21), leptin, insulin-like growth factor-1 (IGF-1) and adiponectin; F) serum β-hydroxybutyrate, acetoacetate, acetone and acetate. Values are β (standard error) derived from linear mixed models with the group, visit and their interaction term (group × time) at 4 and 8 weeks as predictors. The models were adjusted for baseline levels of the outcome and the randomization variable sex. Models with RMR were additionally adjusted for total fat mass. The p-value denotes the difference at 8 weeks. ** data were available for IGF-1 and Adiponectin one baseline and the final visit only.

Baseline characteristics of the participants are presented in **Table 1**. There was an equal number of men (n = 8) in each group, and the mean (SD) age of the participants was 33.2 (5.86) y in the SAAR group and 34.4 (6.26) y in controls. There was a difference of ∼6 kg in baseline body weight between groups (SAAR, mean (SD) 94.4 (10.7); controls, 88.7 (9.22) kg), whereas mean (SD) body fat % was similar (SAAR, 42.0 (6.76); controls, 42.4 (5.80)). All other covariates were comparable between groups.

**Table 1:** Baseline characteristics of the participants in the SAAR and control groups^1^.

|  |  |  |  |  |
| --- | --- | --- | --- | --- |
| **Characteristics** | **SAAR**<br>(n = 31) |  | **Controls**<br>(n = 28) |  |
| Males, n (%) | 8 (25.8) |  | 8 (28.6) |  |
|  | Mean (SD) | Median (IQR) | Mean (SD) | Median (IQR) |
| Age, y | 33.2 (5.86) | 32 (29.5, 37.0) | 34.4 (6.26) | 34 (29.5, 38.8) |
| Body weight, kg | 94.4 (10.7) | 93.2 (86.6, 102) | 88.7 (9.22) | 88.6 (82.0, 95.7) |
| Body mass index, kg/m<sup>2</sup> | 31.4 (2.12) | 31.2 (29.5, 33.2) | 30.8 (2.20) | 30.3 (29.0, 32.4) |
| Waist-to-hip ratio | 0.85 (0.08) | 0.84 (0.79, 0.90) | 0.86 (0.08) | 0.84 (0.79, 0.92) |
| Percent body fat, % | 42.0 (6.76) | 44.0 (37.8, 47.3) | 42.4 (5.80) | 44.1 (39.2, 46.4) |
| Systolic blood pressure, mm/Hg | 120 (10.9) | 119 (111, 128) | 119 (10.7) | 118 (112, 125) |
| Diastolic blood pressure, mm/Hg | 69.1 (7.57) | 70.0 (64.0, 73.0) | 70.0 (8.50) | 66.7 (63.6, 76.1) |
| Apolipoprotein B, g/L | 0.81 (0.20) | 0.80 (0.65, 0.90) | 0.91 (0.24) | 0.90 (0.8, 1.00) |
| Apolipoprotein A1, g/L | 1.49 (0.24) | 1.50 (1.3, 1.6) | 1.47 (0.26) | 1.40 (1.30, 1.55) |
| LDL, mmol/L | 2.81 (0.69) | 2.80 (2.15, 3.35) | 3.11 (0.71) | 3.20 (2.65, 3.50) |
| HDL, mmol/L | 1.40 (0.36) | 1.30 (1.10, 1.65) | 1.39 (0.38) | 1.3 (1.15, 1.50) |
| Total cholesterol, mmol/L | 4.66 (0.75) | 4.90 (4.00, 5.15) | 4.93 (0.83) | 5.00 (4.35, 5.35) |
| Triglycerides, mmol/L | 1.15 (0.55) | 1.00 (0.7, 1.3) | 1.07 (0.42) | 1.00 (0.8, 1.35) |
| Glucose, mmol/L | 5.16 (0.41) | 5.20 (4.90, 5.45) | 5.31 (0.50) | 5.4 (5.05, 5.60) |
| Insulin, pmol/L | 81.0 (47.0) | 77.0 (43.5, 96.0) | 75.5 (39.9) | 66.0 (47.5, 99.5) |
| ASAT, U/L | 24.3 (8.83) | 24.0 (19.0, 26.0) | 22.6 (6.93) | 20.0 (18.0, 25.5) |
| ALAT, U/L | 25.4 (9.64) | 24.0 (20.0, 27.0) | 27.9 (17.3) | 22.0 (17.5, 31.0) |
| Creatinine, μmol/L | 66.4 (11.4) | 67.0 (56.5, 74.5) | 69.0 (11.9) | 72.0 (60.5, 76.5) |
| Resting metabolic rate, kcal/d | 2016 (279) | 2016 (1814, 2131) | 1944 (288) | 1944 (1728, 2145) |
| Respiratory quotient | 0.79 (0.05) | 0.79 (0.76, 0.83) | 0.79 (0.03) | 0.79 (0.77, 0.81) |
| <sup>1</sup>Abbreviations: ALAT, alanine aminotransferase; ASAT; aspartate aminotransferase; HDL; High-density lipoprotein, LDL; low-density lipoprotein; SAAR, sulfur amino acid restriction; All variables are mean (standard deviation) and median (IQR) except row for males. |  |  |  |  |

### 2.2. SAAR reduces urinary sulfate excretion compared to controls

The effects of SAAR on urinary excretion of the compliance biomarker sulfate are presented in **Figure 1B** and **Table 2**. Twenty-four-hour urinary excretion of sulfate decreased in the SAAR vs. control group after 8 weeks (β (95 % confidence interval [CI]) -20.2 (-24.9, -15.6) mol/24 h, p < 0.001).

**Table 2.** Effects of SAAR on body weight, body composition and resting metabolic rate^1^.

|  |  |  |  |  |
| --- | --- | --- | --- | --- |
|  | **4 weeks** |  | **8 weeks** |  |
|  | **β (95% CI)**<br>**SAAR vs. control** | **p-value** | **β (95% CI)**<br>**SAAR vs. control** | **p-value** |
| Body weight, kg | -0.70 (-1.6, 0.21) | 0.13 | -1.14 (-2.04, -0.25) | 0.012 |
| BMI, kg/m<sup>2</sup> | -0.19 (-0.49, 0.12) | 0.23 | -0.30 (-0.60, 0.00) | 0.049 |
| Body fat, % | -0.08 (-0.66, 0.51) | 0.79 | 0.01 (-0.57, 0.59) | 0.98 |
| Total fat, kg | -0.30 (-1.01, 0.42) | 0.41 | -0.40 (-1.11, 0.31) | 0.27 |
| Arm fat, kg | -0.03 (-0.15, 0.09) | 0.67 | -0.02 (-0.14, 0.09) | 0.68 |
| Leg fat, kg | -0.10 (-0.39, 0.19) | 0.50 | -0.15 (-0.44, 0.14) | 0.31 |
| Trunk fat, kg | -0.13 (-0.67, 0.40) | 0.62 | -0.20 (-0.73, 0.33) | 0.46 |
| Android fat, kg | 0.01 (-0.10, 0.13) | 0.80 | -0.05 (-0.17, 0.06) | 0.38 |
| Gynoid fat, kg | -0.03 (-0.16, 0.10) | 0.66 | -0.12 (-0.26, 0.01) | 0.060 |
| Visceral adipose tissue, kg | 0.05 (-0.03, 0.14) | 0.21 | 0.02 (-0.06, 0.11) | 0.57 |
| Subcutaneous adipose tissue, kg | -0.03 (-0.13, 0.07) | 0.55 | -0.08 (-0.17, 0.02) | 0.12 |
| Fat free mass, kg | -0.38 (-0.87, 0.12) | 0.13 | -0.69 (-1.18, -0.20) | 0.013 |
| Resting metabolic rate, kcal/d | 57.1 (-19.4, 134) | 0.14 | 15.2 (-60.4, 90.7) | 0.69 |
| Fat oxidation, g/d | 20.0 (1.82, 38.2) | 0.031 | 12.8 (-5.12, 30.7) | 0.16 |
| Carbohydrate oxidation, g/d | -21.5 (-58.3, 15.3) | 0.25 | -17.9 (-54.2, 18.3) | 0.33 |
| Respiratory quotient | -0.01 (-0.03, 0.01) | 0.20 | -0.01 (-0.03, 0.01) | 0.22 |
| <sup>1</sup> β-estimates and p-values are derived from linear mixed models with body mass as the outcome, and group, visit and their interaction term (group × time) at 4 and 8 weeks as predictors. The models were baseline adjusted. The p-value indicates the difference between groups at each timepoint. Abbreviations: BMI, body mass index; SAAR, sulfur amino acid restriction |  |  |  |  |

### 2.3. SAAR reduces body weight but not resting energy expenditure compared to controls

The effects of SAAR on body weight, body composition, energy expenditure and substrate oxidation are presented in **Figure 1C-D** and **Table 2**.

Both groups lost weight during the intervention. On average, the SAAR group lost 1.14 kg more body weight compared to controls after 8 weeks (β (95 % CI) -1.14 (-2.04, - 0.25) kg, p = 0.012). No conclusive differences between groups were observed for body fat %.

For body composition, BMI and total fat free mass were reduced in the SAAR group compared to controls (**Table 2**). Reductions in gynoid and subcutaneous adipose tissue mass were also observed in the SAAR group compared to controls, but these findings were inconclusive. There were no differences between groups in other regional adipose tissue depots (arm fat, leg fat, trunk fat, android adipose tissue, visceral adipose tissue). Because loss of fat mass and fat free mass directly depend on absolute weight loss, we further adjusted these outcome models for total weight loss. This adjustment attenuated the observed differences between groups, and the effect on fat free mass was no longer statistically significant (data not shown).

Fat free mass-adjusted resting metabolic rate (RMR) was reduced similarly in both groups after 8 weeks. Fat oxidation increased in the SAAR group compared to controls after four weeks (β (95 % CI) 20.0 (1.82, 38.2) g/d, p = 0.031). This effect on fat oxidation remained elevated but was attenuated after eight weeks in SAAR vs. controls (β (95 % CI) 12.8 (-5.12, 30.7) g/d, p = 0.16). No conclusive differences between groups were observed for respiratory quotient (RQ) or glucose oxidation.

### 2.4. SAAR reduces serum leptin and increases plasma ketone bodies

The effects of SAAR on plasma hormones and biomarkers are presented in **Table 3 and Figure 1E-F**. There was a greater decrease of serum leptin in SAAR vs. controls (β (SE) -6.01 (-11.4, -0.59) ng/mL, p = 0.03) after eight weeks, whereas no conclusive effects were observed for other hormones, including FGF-21, IGF-1, adiponectin, and satiety hormones (**Table 3**).

**Table 3.** Effects of SAAR on body weight, body composition and resting metabolic rate^1^.

|  |  |  |  |  |
| --- | --- | --- | --- | --- |
|  | **4 weeks** |  | **8 weeks** |  |
|  | **β (95 % CI)**<br>**SAAR vs. control** | **p-value** | **β (95 % CI)**<br>**SAAR vs. control** | **p-value** |
| ***Urinary sulfur amino acids*** |  |  |  |  |
| Methionine excretion, mmol/24h | -3.84 (-5.96, -1.72) | < 0.001 | -2.70 (-4.86, -0.55) | 0.014 |
| tHcy excretion, mmol/24h | -3.03 (-4.80, -1.26) | < 0.001 | -3.04 (-4.84, -1.24) | < 0.001 |
| Cystathionine excretion, mmol/24h | -30.4 (-46.7, -14.1) | < 0.001 | -32.2 (-48.8, -15.6) | < 0.001 |
| tCys excretion, mmol/24h | -52.7 (-101, -4.66) | 0.03 | -77.3 (-126, -28.4) | < 0.001 |
| Sulfate excretion, mol/24h | -22.3 (-26.9, -17.7) | < 0.001 | -20.2 (-24.9, -15.6) | < 0.001 |
| ***Plasma sulfur amino acids*** |  |  |  |  |
| Plasma methionine, μmol/L | -0.60 (-2.30, 1.11) | 0.49 | -1.26 (-2.94, 0.43) | 0.14 |
| Plasma tHcy, μmol/L | 1.87 (1.01, 2.73) | < 0.001 | 2.07 (1.22, 2.92) | < 0.001 |
| Plasma cystathionine, μmol/L | -0.16 (-0.23, -0.09) | < 0.001 | -0.16 (-0.23, -0.09) | < 0.001 |
| Plasma tCys, μmol/L | 13.0 (-2.17, 28.1) | 0.09 | 12.3 (-2.67, 27.3) | 0.11 |
| ***Plasma biomarkers*** |  |  |  |  |
| Triglycerides, mmol/L | 0.01 (-0.17, 0.20) | 0.90 | -0.13 (-0.32, 0.05) | 0.15 |
| Total cholesterol, mmol/L | 0.05 (-0.26, 0.36) | 0.75 | -0.07 (-0.38, 0.24) | 0.67 |
| LDL, mmol/L | -0.02 (-0.26, 0.23) | 0.90 | -0.05 (-0.29, 0.2) | 0.70 |
| HDL, mmol/L | 0.05 (-0.05, 0.14) | 0.35 | 0.05 (-0.04, 0.15) | 0.29 |
| Apolipoprotein B, g/L | 0.01 (-0.07, 0.09) | 0.78 | -0.01 (-0.09, 0.07) | 0.76 |
| Apolipoprotein A1, g/l | 0.03 (-0.06, 0.11) | 0.52 | -0.01 (-0.09, 0.07) | 0.82 |
| Fasting glucose, mmol/L | 0.00 (-0.19, 0.18) | 0.98 | -0.07 (-0.25, 0.11) | 0.42 |
| ***Plasma/serum hormones*** |  |  |  |  |
| Fasting insulin, pmol/L | -2.24 (-19.3, 14.8) | 0.80 | -9.24 (-26.1, 7.62) | 0.28 |
| C-peptide, pmol/L | 1.49 (-94.8, 97.7) | 0.98 | -59.4 (-155, 35.7) | 0.22 |
| TSH, mU/L | -0.07 (-0.40, 0.27) | 0.70 | -0.30 (-0.63, 0.03) | 0.08 |
| Free T4, pmol/L | 0.08 (-1.17, 1.33) | 0.90 | -0.29 (-1.53, 0.95) | 0.64 |
| FGF-21, pg/mL<sup>2</sup> | -124 (-503, 256) | 0.52 | -83.3 (-460, 294) | 0.66 |
| IGF-1, pg/mL | - |  | -11.4 (-41.7, 18.9) | 0.46 |
| Adiponectin, pg/mL | - |  | -1.43 (-4.43, 1.58) | 0.35 |
| Gastric inhibitory peptide, pg/mL | -11.5 (-68.5, 45.6) | 0.69 | 23.9 (-32.7, 80.4) | 0.41 |
| Glucagon-like peptide 1, pg/mL | -11.7 (-21.3, -2.11) | 0.017 | -7.82 (-17.3, 1.70) | 0.11 |
| Ghrelin, pg/mL | -6.36 (-46.8, 34.0) | 0.76 | -11.7 (-51.8, 28.3) | 0.56 |
| Leptin, ng/mL | -4.06 (-9.53, 1.40) | 0.14 | -6.01 (-11.4, -0.59) | 0.030 |
| Pancreatic peptide, pg/mL | 39.3 (-26.8, 105) | 0.24 | 47.3 (-18.4, 113) | 0.16 |
| Peptide YY, pg/mL | -5.90 (-18.8, 6.99) | 0.37 | -0.69 (-13.5, 12.1) | 0.91 |
| ***Ketone bodies*** |  |  |  |  |
| β-hydroxybutyrate, μmol/L | 46.7 (-54.9, 148) | 0.37 | 117 (16.0, 217) | 0.023 |
| Acetoacetate, μmol/L | 11.2 (-15.0, 37.4) | 0.40 | 27.0 (1.04, 52.9) | 0.042 |
| Acetone, μmol/L | 6.33 (-7.19, 19.9) | 0.39 | 3.67 (-9.71, 17.1) | 0.085 |
| Acetate, μmol/L | 4.28 (-5.45, 14.0) | 0.36 | 8.45 (-1.18, 18.1) | 0.59 |
| ***Safety markers*** |  |  |  |  |
| ASAT, U/L | 3.20 (-0.91, 7.31) | 0.13 | 0.76 (-3.27, 4.79) | 0.71 |
| ALAT, U/L | 1.44 (-5.17, 8.06) | 0.67 | -1.89 (-8.44, 4.65) | 0.57 |
| Urea, mmol/L | -0.27 (-0.63, 0.09) | 0.14 | -0.10 (-0.46, 0.26) | 0.59 |
| Creatinine, $\mu\text{mol/L}$ | 3.82 (1.09, 6.54) | 0.008 | 3.67 (0.97, 6.37) | 0.009 |
| Vitamin B12, pmol/L | 54.5 (-2.86, 112) | 0.06 | 9.31 (-47.4, 66.1) | 0.75 |
| MMA, $\mu\text{mol/L}$ | 0.00 (-0.03, 0.04) | 0.85 | 0.03 (-0.01, 0.06) | 0.17 |

<sup>†</sup> $\beta$ -estimates and p-values are derived from linear mixed models with body mass as the outcome, and group, visit and their interaction term (group × time) at 4 and 8 weeks as predictors. The models were adjusted for baseline values of the outcome.
The p-values indicate the difference between groups at each timepoint.
Abbreviations: ALAT, alanine aminotransferase; ASAT, aspartate amino transferase; BMI, body mass index; FGF-21, fibroblast growth-factor 21; IGF-1, insulin-like growth factor 1; MMA, methylmalonic acid; SAAR, sulfur amino acid restriction; SE, standard error; tCys, total cysteine; tHcy, total homocysteine; TSH, thyroid stimulating hormone

Both groups increased their serum concentrations of ketone bodies (**Figure 1F**, **Table 3**). The increase in plasma concentrations of β-hydroxybutyrate and acetoacetate was greater in SAAR compared to controls, whereas a trend for a greater increase was observed for acetone.

There were no effects of SAAR on classical metabolic risk biomarkers including the plasma/serum lipid profile, glucose metabolism, or any safety markers with the exception of creatinine; a decrease in serum creatinine was observed in the control group, but remained unchanged in the SAAR group (**Table 3**).

### 2.5. SAAR reduces urinary excretion of SAA

The effects of SAAR on urinary excretion and plasma concentrations of SAA are presented in **Figure 2A-B** and **Table 3**. Twenty-four-hour urinary output of methionine, total homocysteine (tHcy), cystathionine, and tCys decreased in SAAR compared to controls (**Figure 2A)**. In plasma, concentrations of tHcy increased and plasma cystathionine decreased in the SAAR group vs. controls (**Figure 2B)**. No conclusive effects were observed for plasma methionine and tCys.

**Figure 2.**
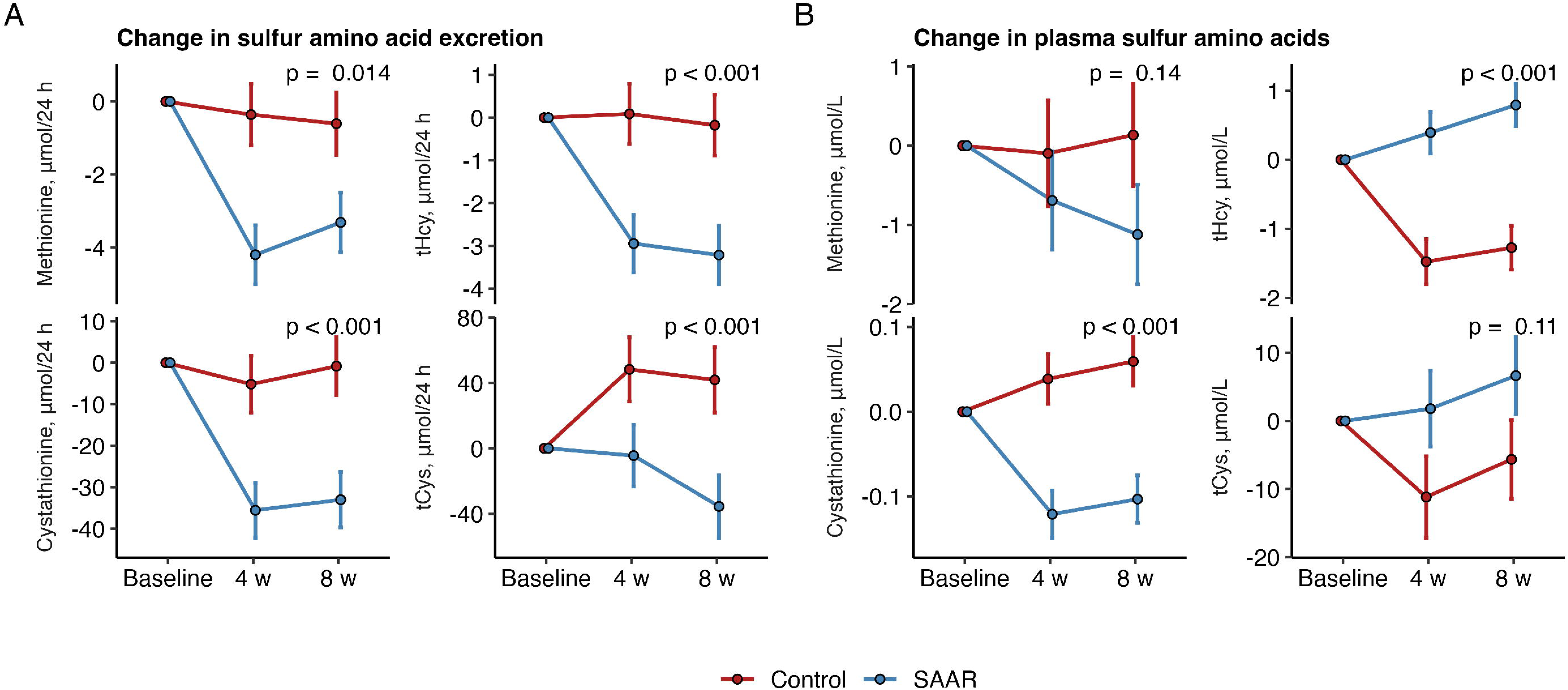
Effects of SAAR on A) sulfur amino acid excretion in 24 h urine and B) change in plasma sulfur amino acids. The models were adjusted for baseline levels of the outcome and the randomization variable sex. The p-value denotes the difference at 8 weeks. Abbreviations: SAAR, sulfur amino acid restriction; tCys, total cysteine; tHcy, total homocysteine. The p-value indicates difference between groups after 8 weeks

### 2.6. SAAR affects adipose tissue gene expression

WAT was available from n = 51 participants at baseline and after eight weeks. In total, 44 genes were differentially expressed at a false discovery rate of 5 % (**Figure 3** and **Table 4**). The full list of genes is available as a supplemental file (**Supplemental File 1**). Twenty-four genes were significantly downregulated including *HSFX2*, *ESM1*, *SIK1B* and *APLN*. Twenty genes were significantly upregulated, including *HIST2H4A*, *ARID5B*, *CYP4B1* and *ADH1B*. The top 10 upregulated and downregulated gene ontologies as identified by generally applicable gene set enrichment (GAGE) analyses are presented in **Figure 3B** and **3C**. GAGE analysis revealed that gene ontologies related to ribosomes were upregulated whereas gene ontologies related to transcription regulation and membrane components were downregulated (FDR < 5 %).

**Figure 3.**
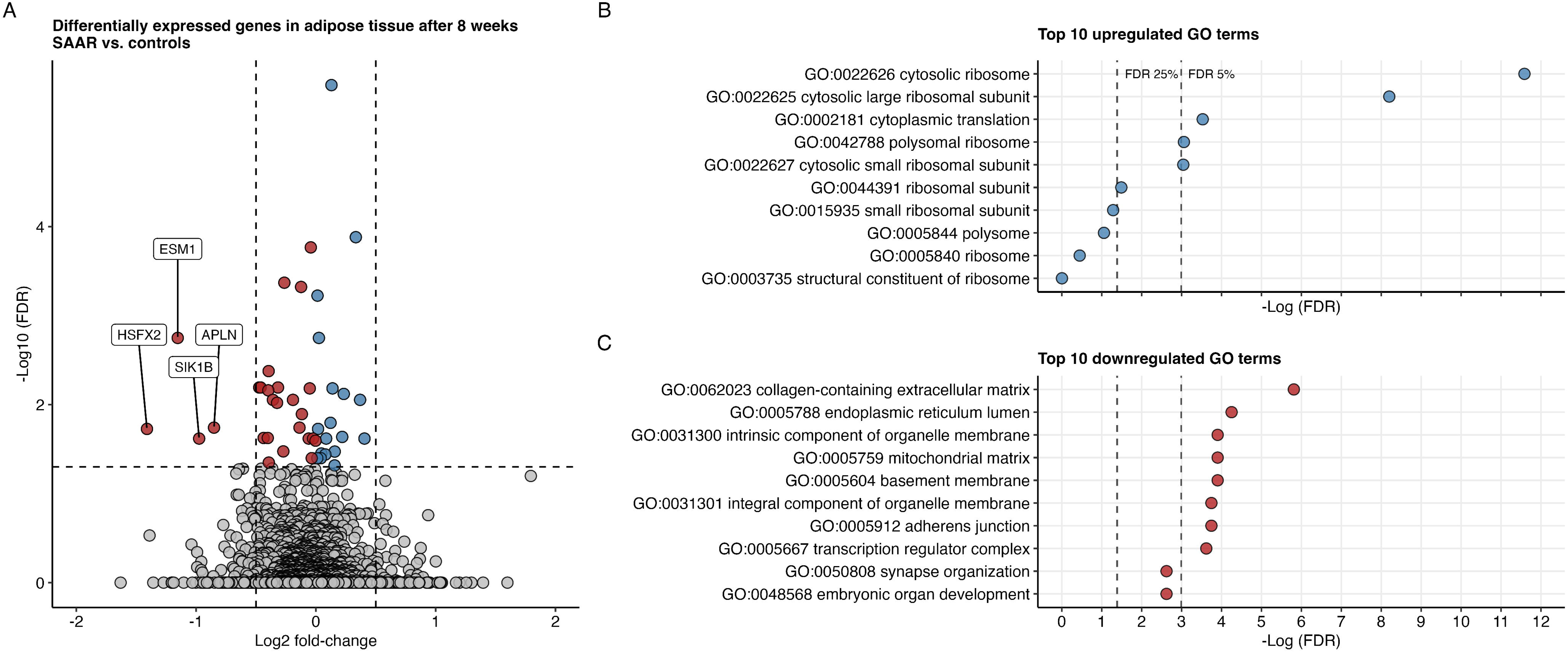
A) Effects of SAAR on adipose tissue gene expression in n = 51 men and women that completed the dietary intervention. Red points indicate negative changes, blue points indicate positive changes. Vertical dashed lines at log2 fold-change of -0.5 and 0.5. Horizontal dashed line indicates FDR of 5 %. Data were available at baseline and after 8 weeks. B) shows the top 10 upregulated gene ontologies and C) shows the top 10 downregulated gene ontologies. Vertical dashed lines denote FDR 25 % and 5 %.

**Table 4:** Significantly differentially expressed genes ranked by Log2 fold-change.

|  |  |  |  |
| --- | --- | --- | --- |
| **Symbol** | **Gene name** | **Log2 fold-change** | **FDR** |
| HSFX2 | heat shock transcription factor family, X-linked 2 | -1.41 | 0.019 |
| ESM1 | endothelial cell specific molecule 1 | -1.15 | 0.002 |
| SIK1B | salt inducible kinase 1B (putative) | -0.97 | 0.024 |
| APLN | apelin | -0.85 | 0.018 |
| LDOC1 | LDOC1 regulator of NFkB signaling | -0.47 | 0.006 |
| DGAT2 | diacylglycerol O-acyltransferase 2 | -0.46 | 0.006 |
| GPAM | glycerol-3-phosphate acyltransferase, mitochondrial | -0.44 | 0.024 |
| DLL4 | delta like canonical Notch ligand 4 | -0.40 | 0.007 |
| MEGF9 | multiple EGF like domains 9 | -0.40 | 0.024 |
| ISYNA1 | inositol-3-phosphate synthase 1 | -0.39 | 0.004 |
| TBX6 | T-box transcription factor 6 | -0.39 | 0.045 |
| CD34 | CD34 molecule | -0.36 | 0.009 |
| ELOVL5 | ELOVL fatty acid elongase 5 | -0.32 | 0.006 |
| RFLNB | refilin B | -0.32 | 0.01 |
| STK40 | serine/threonine kinase 40 | -0.27 | 0.034 |
| SLC35G1 | solute carrier family 35 member G1 | -0.26 | < 0.001 |
| ACBD4 | acyl-CoA binding domain containing 4 | -0.19 | 0.009 |
| ZSWIM7 | zinc finger SWIM-type containing 7 | -0.14 | 0.018 |
| ANLN | anillin, actin binding protein | -0.12 | < 0.001 |
| PPM1N | protein phosphatase, Mg<sup>2+</sup>/Mn<sup>2+</sup> dependent 1N (putative) | -0.12 | 0.013 |
| NOLC1 | nucleolar and coiled-body phosphoprotein 1 | -0.06 | 0.024 |
| SPTA1 | spectrin alpha, erythrocytic 1 | -0.05 | 0.007 |
| KIAA1324 | endosome-lysosome associated apoptosis and autophagy regulator 1 | -0.04 | < 0.001 |
| TBC1D1 | TBC1 domain family member 1 | -0.03 | 0.04 |
| JMJD7-PLA2G4B | JMJD7-PLA2G4B readthrough | -0.02 | 0.024 |
| MAP7D3 | MAP7 domain containing 3 | 0.00 | 0.025 |
| SLC25A24 | solute carrier family 25 member 24 | 0.01 | < 0.001 |
| ITFG1 | integrin alpha FG-GAP repeat containing 1 | 0.01 | 0.04 |
| TEF | TEF transcription factor, PAR bZIP family member | 0.02 | 0.019 |
| NUMA1 | nuclear mitotic apparatus protein 1 | 0.03 | 0.002 |
| LARS2 | leucyl-tRNA synthetase 2, mitochondrial | 0.04 | 0.04 |
| SEMA4A | semaphorin 4A | 0.05 | 0.035 |
| LRRC37A7P | leucine rich repeat containing 37 member A7, pseudogene | 0.08 | 0.036 |
| LRRCC1 | leucine rich repeat and coiled-coil centrosomal protein 1 | 0.09 | 0.024 |
| SETBP1 | SET binding protein 1 | 0.12 | 0.016 |
| TNS1 | tensin 1 | 0.13 | < 0.001 |
| LYG1 | lysozyme g1 | 0.14 | 0.007 |
| TENT5A | terminal nucleotidyltransferase 5A | 0.16 | 0.034 |
| HDAC9 | histone deacetylase 9 | 0.16 | 0.048 |
| TRPS1 | transcriptional repressor GATA binding 1 | 0.22 | 0.023 |
| ADH1B | alcohol dehydrogenase 1B (class I), beta polypeptide | 0.23 | 0.008 |
| CYP4B1 | cytochrome P450 family 4 subfamily B member 1 | 0.33 | < 0.001 |
| ARID5B | AT-rich interaction domain 5B | 0.37 | 0.009 |
| HIST2H4A | novel transcript, antisense HIST2H4A | 0.41 | 0.024 |

Abbreviations: FDR, false discovery rate

## 3. Discussion

### 3.1. Principal findings

In this double-blind randomized controlled dietary intervention study, we aimed to investigate whether effects of SAAR identified in various animal species over the past 30 years could be translated to humans. Our main findings were that dietary SAAR recapitulates some of these benefits in adult humans with overweight and obesity compared to controls, including decreased body weight, BMI, and serum leptin concentrations, and increased plasma concentrations of the ketone bodies β-hydroxybutyrate and acetoacetate^11–24^. In addition, dietary SAAR affected gene expression in WAT.

### 3.2. SAAR, body weight and energy expenditure

We are the first to demonstrate that dietary SAAR with a plant-based diet leads to greater weight loss compared to controls in a double-blind trial with matching diets between the intervention and control groups. This is in contrast to the only other such study with methionine restriction in humans at risk of metabolic disease^33^, and may in part be explained by the lower drop-out rate in the present study, with a key difference being our whole-food approach and provision of the full study diet. The weight loss effect in both groups in the present study is comparable to other dietary interventions with vegan diets^34^, and our study suggests that SAAR may add to this effect. The mechanisms behind this additive effect of SAAR on body weight in humans are unknown. However, despite greater weight loss with SAAR, there were no clear differences in RMR between groups. These findings are partly in line with animal studies, where SAAR increases 24-h energy expenditure despite lower body weights^12,14,23,26,35^. Comparable to the previous study with methionine restriction in humans^33^, we observed a significant increase in fat oxidation after 4 weeks. Although this effect was attenuated after 8 weeks, fat oxidation remained elevated as further supported by the elevated plasma concentrations of ketone bodies in the SAAR group compared to controls. Collectively, the effects of SAAR on body weight may involve regulation of RMR and substrate oxidation, but these effects need to be confirmed in future studies.

### 3.3. SAAR and leptin

Our results are in line with several animal studies showing that SAAR decreased circulating leptin concentrations^12,17,22,23^ and *Lep* expression in adipose tissue^36^. In rodents, decreases in circulating leptin concentrations correlates with decreased visceral fat mass and body weight ^12^. In humans, leptin plays a role in body weight regulation and food intake, and it has been suggested that decreased leptin signaling after weight loss precede adaptive thermogenesis, decreased RMR and increased fat oxidation^37,38^. If so, the decrease in serum leptin in the present study may be a result of the weight loss differences between groups, and not directly due to SAAR. However, one rodent study showed that SAAR, when compared to calorie restriction, reduced mRNA and plasma levels of leptin^14^, indicating that SAAR may have distinct effects on leptin in humans. Notably, there were no effects of SAAR on WAT *LEP* expression in the present study.

### 3.4. Biomarkers of SAA intake

We show for the first time that urinary output of sulfate, an end-product of methionine and cysteine metabolism, decreases significantly with SAAR. Taken together with the strong effects of SAAR on urinary excretion of SAA and related metabolites, the finding on sulfate indicates excellent participant adherence in both groups. The less conclusive effects of diet on plasma methionine and tCys were expected based on similar findings in previous trials and large cohorts^39–44^. The lack of effects on plasma SAA indicate that dietary SAAR activates compensatory mechanisms to maintain availability of particularly cysteine^45^. Indeed, taurine and glutathione metabolism are altered in mice on low-cysteine diets to maintain cysteine availability^46,47^, but whether any such processes are active in humans is not known. Future studies are needed to fully understand the effects of dietary SAAR on circulating and urinary sulfur-containing metabolites in humans.

### 3.5. SAAR and WAT gene expression

This is the first study to demonstrate effects of SAAR on WAT gene expression profiles using a transcriptome-wide approach in humans. Altered gene expression plays a key role in adipose tissue remodeling during SAAR in animals. Among the downregulated genes were *APLN* and *ESM1*, which have been shown to be positively correlated with fat mass^48,49^. Moreover, several genes encoding proteins involved in lipogenesis such as *GPAM*, *ELOVL5* and *DGAT2* were downregulated. The latter finding is in contrast to animal studies, which have reported increased lipogenic gene expression in adipose tissue in conjunction with increased beta-oxidation, and decreased lipogenic gene expression in liver^17^. The reason for this discrepancy is unknown but may be driven by the greater weight loss in the SAAR group. Furthermore, GAGE analyses revealed that SAAR was associated with a seemingly paradoxical regulation of GO terms: SAAR was associated with upregulated cytoplasmic translation, but decreased transcription regulation complex, both of which are implicated in protein synthesis. In animals, SAAR slows overall protein synthesis^50^, but a recent publication outlines a complicated and dynamic machinery in which ribosomal proteins may be both upregulated and downregulated, possibly reflecting a shift in metabolic processes during SAAR^51^. However, these processes were studied in animal livers, and does not necessarily reflect processes in human WAT.

### 3.6. SAAR and fat free mass

In the present study, the SAAR group experienced a greater loss of fat free mass comparison to controls, a finding that is consistent in animal studies with diet-induced obesity^52^. To account for the possibility that this was due to the greater loss of total body weight with SAAR, we created an additional statistical model with adjustment for weight loss. No conclusive effects of SAAR on body mass were found in these models, indicating that the relative loss of fat free mass to loss of total body mass did not differ between the SAAR and control groups. This is further reflected in the lack of observed effects on body fat %. Nevertheless, loss of lean mass is generally not preferable during weight loss, and physical activity should be considered as a co-intervention to maintain lean mass during weight loss with SAAR. It is important to note that we were unable to differentiate between fat free mass compartments (including total body water), and that the predicted loss of approximately 4.3 % of fat free mass in the SAAR group is comparable to other interventions with calorie restriction^53^.

### 3.7. Discrepancies with human and animal studies

A recent controlled feeding study in humans^29^ demonstrated several benefits of SAAR after 4 weeks compared to methionine restriction alone, including reductions in body weight and plasma levels of total cholesterol, LDL, uric acid, leptin, and insulin, blood urea nitrogen, and IGF-1, and increases in body temperature and plasma FGF-21 after 4 weeks. Such effects have also been demonstrated in the animal literature^12–14,22–28^. Although a number of these parameters changed *within* the SAAR group in the present study, there were few differences compared to the control group receiving a diet with identical macronutrient composition with the exception of SAA. Expected phenotypical changes that were not reproduced with SAAR in the present study, included differences in fat mass, improved plasma biomarker profile, increased serum concentrations of FGF-21 and adiponectin, and decreased IGF-1^20,22,24–28,54,55^. These inconsistencies between studies may be due to differences in study designs, control groups, and macronutrient composition. It should also be considered that there are fundamental differences between the composition of dietary SAAR diet used in animals and in humans that may be the reason for the observed discrepancies, limiting the degree of translation across species. Notably, SAAR diets in animals are completely devoid of cysteine, which is impossible to achieve in human diets using whole foods. For example, in rats, cysteine restriction without methionine restriction produced large fluctuations in circulating FGF-21, and cysteine restriction alone appears to be the main driver of the adiponectin response to SAAR^56^. In addition, methionine restriction without cysteine restriction reverses several of the beneficial metabolic effects of SAAR^57^. Therefore, our inability to eliminate cysteine from the diet may partly explain some of the inconsistent results compared to the animal literature. It has additionally been shown that a switch to a vegan diet produces large increases in circulating FGF-21 in humans^58^. Thus, the vegan base diet in both groups in the present study may have obscured the expected SAAR response in serum FGF-21.

Another unexpected result in the present study was the increased serum IGF-1 concentrations in both groups. In the animal literature, decreased IGF-1 signaling is a key mediator of the lifespan-extending effects of SAAR^27,28^. In humans, circulating IGF-1 decreases with protein restriction^59^. However, moderate energy restriction with resulting weight loss, may actually increase circulating concentrations of IGF-1^60,61^, but such results are not consistent^62^. Compared to animal studies where SAAR diets are consumed *ad libitum*, some level of energy restriction was inevitable in our study with fixed energy intakes of ∼2200/2500 kcal/d for women/men in both groups. As noted by others^33^, the effect of weight loss, in both groups, may have obscured potential SAAR-induced effects on circulating IGF-1 and other blood biomarkers. The particular issue with energy restriction and weight loss requires novel approaches to SAAR in humans to capture more of its beneficial metabolic health effects and to increase clinical relevance. Interestingly, a recent study by Plummer & Johnson demonstrated that intermittent SAAR reproduced most, if not all benefits of continuous SAAR^24^. Intermittent SAAR presents an attractive approach for future clinical trials in humans, in which SAAR could be applied on alternating days, reducing the burden on each study participant.

### 3.8. Clinical implications

The implications of a weight loss difference of about ∼1.14 after 8 weeks is likely to be relatively modest, and it is not certain if this effect would persist over the longer term. However, an ∼20 % greater weight loss with SAAR may be an appealing implication for individuals wanting to lose weight. The added possibility that RMR is maintained despite greater weight loss during SAAR, is also an attractive premise for prescription of long-term SAAR-induced weight loss *if* the observed RMR effect persists. Although we have shown that it is possible to implement an 8-week dietary intervention with SAAR with excellent adherence, its feasibility in a setting without close follow-up or full food provision is unknown. Other approaches to limit intake of SAA should therefore be explored.

### 3.9. Strengths and limitations

This study has several strengths, including the randomized and double-blind design, and the broad approach to data collection to thoroughly describe the effect of SAAR in humans. By taking measures against low participant adherence, we were able to maintain a low drop-out rate (10 % vs. expected 25 %) and high compliance, likely due to the close follow-up by registered dietitians and provision of all foods free of charge for the duration of the intervention. Good overall adherence was further indicated by the distinct differences in urinary excretion of sulfate, the end-product of sulfur amino acid oxidation.

Several limitations should be considered; because the study was performed in free-living subjects, we were unable to standardize the intake of SAA to kilogram of body weight. This means that although we succeeded in comparing sharply contrasting absolute levels of the SAA in the diet, intake relative to each subjects’ body weight varied, which likely resulted in increased variation and decreased statistical power. It is reasonable to assume that adjustments for baseline body weight and within-subject correlation in the statistical models handle a portion of this probable source of bias, but other impacts cannot be excluded. Therefore, inpatient studies, where total control of dietary intake and protocol adherence are simpler to implement, would provide more precise information on the biological effects of SAAR in humans, but would do so at the expense of generalizability to the real-life situation. An additional limitation with the present study includes the relatively short study duration of 8 weeks. Lastly, the intake of SAA in the control group may be higher than a typical Western diet, however, relative to body weight the content it is in line with reported intake in upper quantile groups in large population studies including NHANES and the Framingham cohort studies^6–8^.

## 4. Conclusion

In conclusion, a plant-based whole food SAAR intervention for 8 weeks led to greater weight loss and decreased serum leptin concentrations and increased plasma concentrations of ketone bodies. In addition, SAAR affected adipose tissue gene expression related to ribosomal proteins, transcription regulation and membrane components. Despite greater weight loss there were no clear differences in RMR between the SAAR and control groups, indicating that SAAR may maintain RMR through mechanisms that may not include typical mediators identified from animal studies. It is not known whether these effects persist over the longer term, and studies with a longer duration should be performed.

## 5. Methods

### 5.1. Study design

Details about study design, recruitment strategies, inclusion and exclusion criteria, randomization, blinding, diet run-in period, dietary intervention, and measures taken to increase adherence can be found in the published study protocol^63^, with additional information in the online **Supplement**.

Briefly, we conducted an 8-week, double-blind, randomized controlled dietary intervention study at the Centre for Clinical Nutrition at the University of Oslo, Norway, from March 2021 to March 2022. Healthy men and women aged 18 to 45 years, with a body mass index (BMI) between 27 and 35 kg/m^2^, were invited to participate. Of the 158 people assessed for eligibility, 72 were invited to participate in the study. Sixty-one participants (45 women, 16 men) accepted the invitation and were randomized in blocks stratified by sex to either the SAAR or the control group with 1:1 allocation ratio. Two participants were unable to start the intervention due to Covid-19 (n = 1) and other illness (n = 1) related reasons, and therefore a total of 59 started the intervention at baseline. All participants gave informed consent and the study was conducted according to the Declaration of Helsinki. The study was approved by the Regional Ethical Committee South-East (Ref no. 123644).

All participants performed a 7-day run-in period with instructions to follow the Nordic Nutrition Recommendations^64^, and to maintain their habitual physical activity. Participants were instructed to fast for 12 h, and to avoid strenuous physical activity, alcohol and caffeine for 24 h prior to the study visit, in accordance with the protocol for RMR measurements in the whole room indirect calorimeter (WRIC).

To blind participants to the intervention, we developed a SAA restricted base diet that was identical in both groups and consisted of vegan plant-based whole foods, as methionine and cysteine are abundant in animal-derived protein and in certain fruit, grains, nuts, and vegetables. In addition, the diet was supplemented with an amino acid powder drink mix without methionine and cysteine (XMET XCYS Maxamaid, Nutricia Norway AS, Oslo, Norway), 10 µg /d of vitamin D (Løft, Sweden) and 225 µg/d of iodine (Nycoplus, Norway) to achieve adequate nutrient intakes in line with the Nordic Nutritional Recommendations^64^. All foods were delivered home to the participant on a weekly basis. Participants received two 7-day menus with detailed recipes that rotated every 2 weeks. To not place excessive limitations on the participants’ social life, they were allowed to replace one regular meal with one *ad libitum* meal per week to increase compliance (see below). In addition, participants were allowed to consume foods without SAA from a pre-specified list *ad libitum*. The mean energy content of the base diet was 2401 kcal/day for men and 2030 kcal/day for women. The relative contribution of total fat, carbohydrates, and protein in the SAAR base diet was approximately 34 energy % (E%), 46 E% and 13 E%, respectively. The mean daily dietary composition with and without an estimated *ad libitum* meal is shown in **Supplemental Table S1.**

The contrasting SAA content between the SAAR and control group was adjusted with capsules. The SAAR group was given 8 capsules containing a total of 3 g/day of maltodextrin, whereas the control group was given 8 capsules containing a total of 1.125 g/day of methionine and 2.50 g/d of cysteine, and were recommended to distribute these evenly with meals. In the SAAR group, mean intake of total SAA from diet and capsules was 1.9 g/d for men and 1.8 g/d for women. In the control group, mean intake of total SAA from diet and capsules was 5.5 g/day for men and 5.4 g/day for women. This translated to a mean (SD) intake of SAA of 19.2 (1.83) g/kg/d in SAAR and 62.1 (6.07) g/kg/d in controls. The intake of SAA in the SAAR and the control groups are in line with the FAO recommended daily allowance of 19 mg/kg/d^65^ and the median in the upper intake quintile of SAA in the National Health and Nutrition Examination Survey (NHANES) (62.7 mg/kg/d)^6^, respectively. Examples of daily meals in the base diet are given in **Supplemental Table S2**.

To maintain participant adherence, the participants received all foods free of charge delivered to their homes once weekly. Furthermore, two follow-up conversations were scheduled with registered dietitians, whom they could otherwise contact at any time during the study period if necessary. In addition, the allowance of one self-chosen *ad libitum* meal per week was included to limit consequences for their social life. The composition of this meal was recorded in a food diary.

Subjective compliance to the dietary interventions were recorded using food diaries and evaluation forms, and the participants were required to return any capsules that were not consumed. Objective markers of compliance included 24-h excretion of sulfate, methionine total homocysteine (tHcy), cystathionine, and tCys. The primary outcome in the present study was body weight. Because the power calculation was only done for body weight and not secondary outcomes, the other outcomes were regarded as exploratory. The latter outcomes included SAA profile in plasma (methionine, tHcy, cystathionine, and tCys) and 24-h urine (with the addition of sulfate); fat mass (total, arm, leg, trunk, gynoid, android, visceral and subcutaneous); fat free mass; RMR and its components (respiratory quotient [RQ], fat oxidation, glucose oxidation); adipose tissue gene expression profiles; blood biomarkers related to metabolic health (total cholesterol, low-density cholesterol, high-density cholesterol, apolipoprotein B, apolipoprotein A1, triglycerides, insulin, glucose, C-peptide); endocrine factors (FGF-21, IGF-1, adiponectin, leptin, thyroid-stimulating hormone, Free T4, insulin, C-peptide, gastric inhibitory peptide-1, glucagon-like protein-1, peptide YY, pancreatic peptide, ghrelin); plasma ketone bodies (β-hydroxybutyrate, acetoacetate, acetone, acetate); and other/safety markers (vitamin B12, methylmalonic acid, creatinine, urea, alanine aminotransferase [ALAT] and aspartate aminotransferase [ASAT]). Details of outcome assessment are given below and in the online **Supplement**. All outcomes were measured at baseline, 4 and 8 weeks, except for IGF-1 and adiponectin (baseline and 8 weeks).

### 5.2. Anthropometry and body composition

Body anthropometry (waist and hip circumference, height and weight) were measured according to standardized operating procedures with the participant wearing light clothing. Waist and hip circumference were measured thrice and the average was used. Standing height and weight were measured once using the electronic height board (SECA 285, SECA, Germany). All measurements were recorded with one decimal. Body composition was measured by the Dual-energy X-ray absorptiometry (DXA) procedure (Lunar iDXA, GE Healthcare Lunar, Buckinghamshire, United Kingdom). The software enCORE ™ version 18 was used for data acquisition. DXA scans were performed on fasting subjects using standardized procedures adapted from the International Society for Clinical Densitometry^66^.

### 5.3. Blood pressure measurements

Blood pressure was measured using a CARESCAPE V100 Vital Signs Monitor (GE Healthcare, Chicago, Illinois, USA) with the participant in a seated position and the average of three readings was used.

### 5.4. Whole-room indirect calorimetry

Resting metabolic rate (RMR; kcal/d), macro-nutrient oxidation (g/d) and the RQ [liters (V) of carbon dioxide (CO_2_)/liters of oxygen (O_2_)] were measured using a whole-room indirect calorimeter (WRIC) specific for RMR. This involved the determination of O_2_ (%), CO_2_ (%) and water vapor pressure (WVP, kPa) according to previously published protocols ^67,68^.

After arriving to the research center in the morning, subjects urinated and were placed inside the RMR-WRIC in a comfortable recliner for 60 minutes. They were instructed to remain seated, minimize movements and keep the lights on. To prevent subjects from falling asleep, they were encouraged to bring a book or an iPad to watch TV.

Metabolic rate (kcal/min) was calculated using Weir’s equation^69^ along with the RQ, fat oxidation and glucose oxidation. Substrate oxidation was calculated as published previously ^70^. Mean RMR (kcal/min), fat oxidation (g/min) and glucose oxidation (g/min) were multiplied by 1440 to convert to kcal/day or g/day. The WRIC measurements have been validated against 10 propane combustion tests^67^ with a mean (SD) error of -1.3 (1.1) % for RMR and -1.4 (0.5) % for RQ and repeated measurements in 19 eligible subjects with a within-person coefficient of variation of 2.68 %^71^.

### 5.5. Blood samples

Fasting venous blood samples were collected at each study visit, and included one 4 mL EDTA-lined vacuum tube, one lithium-heparin 5 mL tube, and one serum gel 5 mL for immediate analyses of routine markers. One additional lithium-heparin 5 mL tube for amino acid assays including methionine, tHcy, cystathionine, and tCys was collected. One additional 5 mL serum gel tube was collected for analysis of adipokines and hepatokines.

All samples were inverted 6-8 times directly after withdrawal. For immediate analyses of routine markers, the 4 mL EDTA tube was stored at 4 °C whereas the 5 mL lithium-heparin tube was centrifuged at 2500 *g* at 4 °C for 10 min, and stored at 4 °C. The 5 mL serum tubes were left in room temperature for a minimum of 30 min to ensure clot activation and then centrifuged for 10 min at 1500 *g*. Serum was stored at –20 °C until analysis of routine markers. For amino acid assays, one 5 mL lithium-heparin tube for amino acid assays was directly placed in ice water, and centrifuged within 15 min at 2000 *g* at 4 °C for 5 min. Immediately after centrifugation, a minimum of 650 µl were transferred to cryotubes and stored at – 80 °C until analysis. Amino acid assays were performed at the Department of Pediatrics and Inherited Metabolic Disorders, First Faculty of Medicine, Charles University and General University Hospital in Prague. The samples were shipped on dry ice.

### 5.6. Urine samples

A 24-h urine collection took place before each study visit and started after the participants had emptied their bladder for the first time the morning prior. All urine after this point was collected in 3 L containers with light protection and stored at 4°C. Twenty-four-hour urine were subsequently aliquoted and stored at – 80°C until analysis, and shipped on dry ice to Prague.

### 5.7. Biochemical analyses

Sulfur-containing compounds except sulfate were quantified as described elsewhere ^72^. Briefly, tCys and tHcy were assayed by high-performance liquid chromatography after reduction with Tris(2-carboxyethyl)phosphine and derivatization with 7-fluorobenzo-2-oxa-1,3-diazole-4-sulfonate while methionine and cystathionine were assayed by liquid chromatography tandem mass spectrometry using a commercial kit from Phenomenex. Sulfate in urine was determined by a newly developed method (Barvíková et al, unpublished) as follows: samples of urine were diluted 100-times with deionized water, calibration standards of sulfate (0-20 mg/L) were prepared by diluting the Multi Anion Standard (certified TraceCERT®, Sigma Aldrich) with deionized water. Inorganic anions were separated on Shodex IC-90 4E column (Showa Denko Europe GmbH, Munich, Germany) with isocratic elution (1.8mM sodium carbonate and 1.7mM sodium bicarbonate) using Ion Exchange Chromatograph with suppressor from Shimadzu (Shimadzu Europa GmbH). Pumps SIL-20Ai and autosampler SIL-20AC were coupled with CDD-10A conductivity detector, the system was coupled with the ion suppressor Xenoic ® Asurex-A200 (Xenoic ® Asurex-AR1 cartridge, Diduco AB, Umeå, Sweden). Urinary sulfate concentration was calculated from calibration curves, converted to mM and expressed as mmol/24 h. All other urinary metabolites were expressed as µmol/24 h.

Routine clinical biomarkers (ASAT, ALAT; creatinine (serum and urine), urea, glucose, insulin, c-peptide, total cholesterol, LDL, HDL, apolipoprotein B and apolipoprotein A1, vitamin B12 and methylmalonic acid) were measured at the Department of Medical Biochemistry (Oslo University Hospital Rikshospitalet, Oslo, Norway) by colorimetric and/or enzymatic methods (Cobas c702 analyzer, Roche Diagnostics International Ltd, Rotkreuz, Switzerland).

Serum concentrations of FGF-21, gastric inhibitory peptide-1, glucagon-like peptide-1, ghrelin, pancreatic peptide, peptide YY and leptin (MSD U-Plex Metabolic Group 1 (Human) - Multiplex Assays Kit Cat No: K151ACL-1), IGF-1 (R&D Systems Human IGF-1 Quantikine ELISA Kit Cat No: DG100B), and adiponectin (MSD R-Plex Human Adiponectin Assay Kit Cat no: K151YTR-2) were measured at AS VITAS using a multiplexed ELISA platform.

Plasma β-hydroxybutyrate, acetoacetate, acetone and acetate were measured using a commercial high-throughput proton NMR metabolomics platform (Nighintgale Health Ltd. Helsinki, Fin) as described previously^73^.

### 5.8. Adipose tissue biopsies

WAT biopsies were available from the first and last study visit and was taken from the periumbilical region. The skin was sterilized, and the skin and subcutis were anaesthetized by injecting 5 mL of a local anesthetic (Xylocaine 10 mg/mL AstraZeneca, Södertälje, Sweden). Subcutaneous WAT were immediately dissected on an ice-cold aluminum plate to remove blood and other materials before aliquots were snap-frozen in 1.5 mL cryotubes and stored at − 80 °C until analysis.

### 5.9. RNA isolation

WAT RNA was isolated using a modified version of the protocol provided by manufacturer (Nucleospin RNA, Mini kit for RNA purification, Macherey-Nagel, Germany). Briefly, approximately 100 mg tissue was homogenized using 750 μl Triazol and centrifuged at 12 000 RCF for 15 minutes at 4 ⍰. Then, 0.3 ml chloroform was added to the sample prior to another centrifugation step at 12 000 RCF for at 15 minutes at 4 ⍰. RNA-containing aqueous phase was extracted and mixed with 96 % EtOH at room temperature. The subsequent steps followed the manufacturer protocol. RNA integrity was measured by the RNA 6000 Nano Kit and an RNA integrity number of > 7 was considered sufficient quality.

### 5.10. mRNA sequencing and bioinformatics

Fifty-one participants had full sets of WAT that were sent to the Norwegian Sequencing Centre (www.sequencing.uio.no) for analysis.

Libraries were prepared with the Illumina TrueSeq stranded mRNA kit using 500 ng input per sample, following the manufacturer’s protocol. Sequencing was performed on an Illumina NovaSeq over two quarters of an S4 flow cell with the XP workflow and 151 bp paired end reads, according to manufacturer’s instructions. Base calling was performed with RTA v3.4.4, and demultiplexing with bcl2fastq v2.20.0.422. Reads quality filtering were performed using BBMap v38.79-GCC-8.3.0. Read T overhang was filtered by removing the 151st base on each read, resulting in 150 bp reads, followed by filtering of adaptor sequences. Read alignment was performed using the ultra-sensitive spliced-aware aligner STAR v2.7.9a-GCC-11.2.0^74^ and pseudo-alignment was performed using Kallisto v0.46.1, both against the GENCODE human genome release 41(GRCh38.p13) primary assembly, including the corresponding annotation file. The union-exon based approach was used for gene quantification using STAR^75^ and the resulting gene counts were normalized using the trimmed mean of the M-values (TMM) algorithm. Transcript level quantification using Kallisto was normalized as transcripts per million (TPM). Subsequent batch correction (a subset of samples was re-sequenced in a separate run) was done using the removeBatchEffect function in edgeR v3.38.4^76^. Down-stream gene level differential expression analysis was performed using DEseq2 v1.38.2, and transcript level differential expression analysis was performed using Sleuth v30.1. P-values were corrected for multiple testing using the Bejamini-Hochberg method for controlling the false discovery rate (FDR) at 5 % ^77^. GAGE was performed using the GAGE v2.44.0 R package for global analyses of regulated pathways. The default native workflow was followed as recommended for mRNA sequencing data ^78^.

### 5.11. Sample size calculations

Sample size calculations have been described in the previously published protocol^63^. Briefly, we hypothesized a -1 kg difference after 4 weeks and -3 kg difference in body weight after 8 weeks between the SAAR group and controls. We hypothesized a correlation of 0.9 for body weight between study visits and a decay rate of 0.5. The online tool GLIMMPSE was used for power calculations (https://v2.glimmpse.samplesizeshop.org). The estimated sample size was 46, and accounting for 25 % drop-out (46/1 - 0.25) the final number was 61.

### 5.12. Statistical analyses

Continuous and categorical variables are presented descriptively as mean (SD) and median (interquartile range [IQR]), and n (%), respectively. No significance test for group differences in participant characteristics (**Table 1)** have been performed in accordance with CONSORT guidelines^79^. As specified in the protocol^63^, blinded outcome analyses were performed by using linear mixed model regression, which is robust to missing values. To correct for any baseline differences, we used the constrained approach as described by Coffman *et al.*^80^ and Twisk^81^. This approach increases precision by forcing baseline values of the outcome to be equal between groups. Thus, the model included the visit covariate, interaction terms for visit and the grouping covariate at 4 weeks and 8 weeks, the stratification variable sex, and a random intercept to control for within-subject correlation. This type of baseline adjustment was performed for all outcome models. The resulting p-values from the interaction terms indicate the difference between groups at each visit (4 weeks and 8 weeks). Unless otherwise specified, model output is presented as β and 95 % CIs. EMMs and corresponding 95 % CIs derived from the models are presented in the online supplement together with the observed values at each timepoint. The main analyses were performed on the ITT population, but PP analyses (in which drop-outs are excluded) are presented in the online supplement. Because we were interested in the direct effect of the diet on body composition independently of total mass loss, additional models for body composition outcomes were created with adjustment for total weight loss. In models with RMR as the outcome, models were adjusted for fat free mass. The assumption of normally distributed model residuals was inspected visually. In initial analyses, residuals for β-hydroxybutyrate, acetoacetate and FGF-21 were severely skewed, and these markers were therefore log-transformed before analysis. A p-value < 0.05 was considered statistically significant. Considering that we did not power the trial for secondary outcomes, all other outcomes are regarded as exploratory and no multiplicity adjustments were performed. All statistical analyses and plots were performed/made in R version 4.2.1 in the R Studio IDE version 2022.02.3 Build 492, using packages in the *tidyverse*, *lmerTest* and *stats*.

## 6. Data availability

Raw data from this study cannot be made publicly available due to existing European privacy regulations. Requests for data can be made to the corresponding author and will be shared pending approved protocols by relevant ethical committees and privacy regulation authorities.

## 7. Author contributions

TO, HR, KJV and KR contributed to study design; BØ developed and designed the base diet for the intervention; TO, ES, HJ, BD, AADV, AFD KS and KJV collected data; ES: had main responsibility for supervision of study participants; TO, ET, SL and MT performed statistical analyses; KB and VK were responsible for biochemical analyses of sulfur amino acids and sulfate; RR developed the protocol for calorimetry measurements and data analysis of raw calorimetry data; AE and MvG contributed intellectually; TO, ES and KJV drafted the manuscript; All authors revised the draft and approved the final version of the manuscript.

## Conflicts of interest

None declared

## 8. Acknowledgements

We wish to thank Elin Augestad for assistance with blood sampling; Elisabeth Andrea Johnsen and Stine Sofie Strømland for assistance with sample handling; and chef Johannes Grødal for developing meals for the dietary intervention. Finally, we wish to thank all study subjects for their time. This project has received funding from The Research Council of Norway (Grant no: 310475) under the umbrella of the European Joint Programming Initiative “A Healthy Diet for a Healthy Life” (JPI HDHL) and of the ERA-NET Cofund HDHL INTIMIC (GA N° 727565 of the EU Horizon 2020 Research and Innovation Programme), and Institute of Basic Medical Sciences, University of Oslo, and Henning och Johan Throne-Holsts stiftelse. VK and KB were supported by the Czech Ministry of Education, Sports and Youth (STAY 8F20013), institutional support was provided by Charles University (program COOPERATIO-Metabolic Disorders) and Ministry of Health DRO VFN64165.

## Notes

### Competing Interest Statement

The authors have declared no competing interest.

### Clinical Trial

NCT04701346

### Clinical Protocols

https://translational-medicine.biomedcentral.com/articles/10.1186/s12967-021-02824-3

### Author Declarations

The Regional Ethical Committee South-East, Norway, gave ethical approval for this work (Ref no. 123644).

