## Supplementary material for "Dietary sulfur amino acid restriction in humans with overweight and obesity: a translational randomized controlled trial": Online supplement

^a^ Shared authorship.

### 1. Supplemental methodological information

#### 1.1. Inclusion and exclusion criteria

Men and women aged 18 to 45 years with a body mass index (BMI) between 27 and 35 kg/m^2^ who were otherwise healthy were invited to participate. We asked individuals with a self-reported BMI of 27 to 30 kg/m^2^ in the screening questionnaire to report waist circumference to confirm traditional cut-offs at > 80 cm for women and > 94 cm for men. Exclusion criteria were chronic use of medication or chronic disease, veganism (> 1 month), smoking, established co-morbidities, pregnancy or breastfeeding the last three months, significant weight loss (> 5 %) over the last three months, and high-intensity training (e.g., interval running, CrossFit, heavy strength training) more than three times per week.

#### 1.2. Recruitment

We recruited participants through the social media channels of the Department of Nutrition, University of Oslo. Of the 158 people assessed for eligibility, 72 were invited to participate in the intervention trial. Sixty-one healthy women and men accepted the invitation and were randomized to either the SAAR group or the control group, and 59 participants started at baseline as two dropped out pre-baseline due to Covid-19-related issues.

#### 1.3. Randomization

Participants were randomized stratified by sex in random block sizes to ensure similar sex distribution and equal sized groups. Randomization was performed by a researcher unrelated to the trial using the *blockrand* package in R (R for Statistical Computing, Vienna, Austria).

#### 1.4. Dietary information

All meals in the dietary intervention were based on a previously developed diet ^1,2^ and recipes were tailored by a professional chef to ensure diverse and tasty dishes. Participants were permitted to drink unlimited amounts of black coffee, tea, water, and diet sodas. Additionally, they were given a list of low SAA-containing foods for extra or *ad libitum* consumption and a list of ingredients to add taste to the dishes. Participants were given flavor enhancers (Flavour Modjul® from Nutricia or an alternative enhancer selected from a pre-defined list) to mix with the powdered drink to mask the unpleasant taste. Diet composition and example meals are presented in **Supplemental Table 1** and **2.**

#### 1.5. Participant adherence

Several measures were taken to maintain participant adherence to their diets. All participants received weekly grocery delivery, containing all foods for the dietary intervention including meals and snacks for the duration of the study. Follow-up conversations by trained registered dietitians or dietitian students in their final year of training, were completed between visits, using telephone calls, SMS, and e-mails. Follow-up conversations were scheduled for the end of week 2 and week 6, where participants were asked questions about diet compliance, and maintenance of the intake of powder, capsules, and vitamins, clinical symptoms, motivation, and if they had any questions. Outside the scheduled follow-up conversations, study participants could contact the registered dietitian at any time. Both subjective and objective data on compliance were collected. Participants were required to fill out a 4-day food diary (3 weekdays, 1 weekend day) prior to each study visit indicating if they had followed the diet or not, and whether they had made use of the *ad libitum* meal. At the final study visit, participants were required to fill out a questionnaire indicating the overall compliance over eight weeks. They were also required to return any capsules that had not been consumed. The main objective markers of compliance were 24 h urinary excretion methionine, total homocysteine, cystathionine, total cysteine and sulfate. We also included plasma concentrations of these metabolites, although they are not as responsive to dietary intake as we have observed under various conditions previously (with the exception of plasma cystathionine) ^1–3^.

#### 1.6. Whole-room indirect calorimetry

Resting metabolic rate (RMR; kcal/d), macro-nutrient oxidation (g/d) and the respiratory quotient (RQ; liters (V) of CO_2_/liters of O_2_) were measured using a whole-room indirect calorimeter (WRIC) specific for resting metabolic rate. This involved the determination of oxygen (O_2_, %), carbon dioxide (CO_2_, %) and water vapor pressure (WVP, kPa) according to previously published protocols ^4,5^. The RMR-WRIC has an interior volume of 7,500 liters, after accounting for the space taken up by the recliner, toilet and sink. Fresh air from outside is infused into the buffer spaces surrounding the WRIC’s by the lab’s ceiling mounted HVAC unit. This HVAC unit assures a slight positive pressure within the entire lab space to prevent any outward air leaks from the WRIC’s. Thereafter, it is heated or cooled by the buildings central HVAC system to maintain a temperature of approximately 22 C. The heat pump unit within the RMR-WRIC maintains a temperature of 22 C and is set to have the fan on continuously at a low setting to mix the respiratory gases (LG D09TR NSJ, LG Electronics, Seoul, South Korea). Excurrent (sample gas from within the RMR-WRIC) air is drawn continuously from the RMR-WRIC at a constant flow rate of 100 L/min and analyzed using the Promethion GA-3m2/FG-250 (Sable Systems International, Las Vegas, NV, US) integrated metabolic instrumentation. This instrumentation contains dual CO_2_, O_2_ and WVP sensors that alternate between measuring gas concentrations in the incurrent (baseline air from the buffer space) and excurrent air streams thus maintaining a continuous measurement of the subject’s respiratory exchange within the RMR-WRIC.

After arriving to the lab in the morning, subjects voided and were placed inside the RMR-WRIC in a comfortable recliner for 60 minutes. They were instructed to remain seated, minimize movements and keep the lights on. To prevent subjects from falling asleep, they were encouraged to bring a book or an iPad to watch TV.

Raw data for O_2_, CO_2_ and WVP were obtained by the Caloscreen acquisition software (Sable Systems International, North Las Vegas, USA, version 1.3.16). Metabolic calculations were performed utilizing ExpeData (Sable Systems International, North Las Vegas, USA, version 1.9.27) utilizing previously published formulas ^6^. Energy expenditure (kcal/min) was then calculated using Weir’s equation ^7^ along with the RQ. RQ was calculated as VCO_2_/VO_2_. Mean RMR (kcal/min) was multiplied by 1440 to convert to kcal/day. The WRIC measurements have been validated against 10 propane combustion^4^ tests with a mean (SD) error of -1.3 (1.1) % for RMR and -1.4 (0.5) % for RQ and repeated measurements in 19 eligible subjects with a within-person CV of 2.68^8^.

### 2. Supplementary tables

#### 2.1. Diet composition and meal examples

| Supplemental Table S1. Mean daily intake* of energy and nutrients from base diet by sex. | | | | |
| --- | --- | --- | --- | --- |
|  | **Men** | | **Women** | |
| Includes *ad libitum* meal estimate^†^ | No | Yes^†^ | No | Yes^†^ |
| Energy, kcal | 2401 | 2564 | 2030 | 2193 |
| Total fat, % of energy | 33.0 | 34.0 | 33.8 | 35.2 |
| Total fat, g | 90.7 | 97.0 | 79.6 | 85.9 |
| Carbohydrates, % of energy | 46.4 | 45.2 | 46.4 | 44.7 |
| Carbohydrates, g | 274 | 290 | 230 | 245 |
| Protein, % of energy | 14.0 | 14.0 | 13.3 | 13.4 |
| Protein, g^1^ | 83.1 | 90.0 | 66.5 | 73.4 |
| Sulfur amino acids, g | 1.9 | 2.1 | 1.8 | 2.0 |
| Methionine, g | 0.9 | 1.1 | 0.8 | 1.0 |
| Cysteine, g | 1.0 | 1.1 | 0.9 | 1.0 |
| Total sulfur amino acids, g, SAAR/control | 1.9/5.5 | 2.1/5.7 | 1.8/5.4 | 2.0/5.6 |
| ^*^Data are expressed as means based on a 2-week menu.  ^†^Participants were allowed one ad libitum meal per week. The present mean daily intake includes an estimate of this meal based on typical social/weekend meals.  SAAR: Sulfur amino acid restriction.  ^1^18.75 g protein was provided by the XCYS XMET supplement for men, and 12.5 g for women. | | | | |

| Supplemental Table S2. Examples of meals in the base diet. | |
| --- | --- |
| **Meals** | **Examples of dishes** |
| Breakfast | Oatmeal with apple. |
|  | Bread rolls with jam/avocado. |
| Lunch | Salad with beans, lentils and/or chickpeas. |
|  | Norwegian vegetable hash with lentils. |
|  | Falafel sandwich. |
|  | Baked broccoli salad with bean dressing. |
|  | Greek salad with vegan tzatziki. |
|  | Tomato salad with bread rolls. |
|  | Fried rice with beans. |
| Dinner | Vegetable casserole or soup. |
|  | Curry-lentil casserole. |
|  | Chili sin carne with rice and salad. |
|  | Falafels with salad and vegan aioli. |
|  | Spiced chickpeas. |
|  | Vegan curry with sweet potatoes, chickpeas, and apples. |
| Supper | Bean and chick-pea salad. |
|  | Broccoli soup with bread rolls. |
|  | Bread rolls with jam/avocado/tomatoes/vegan aioli/banana. |
| Snacks | Fruits and Brazil nuts. |
|  | Kale chips. |
| Powder drink mix | 3-4 amino acid powder free of methionine and cysteine was consumed as drinks throughout the day. |
| Supplements | One vitamin D capsule and one iodine capsule. |

#### 2.2. Intention-to-treat analyses

| Supplemental Table S3. Mean (SD) observed values of all outcomes in the SAAR and control groups^1^. | | | | | | |
| --- | --- | --- | --- | --- | --- | --- |
|  | **Control** | **SAAR** | **Control** | **SAAR** | **Control** | **SAAR** |
|  | **Baseline** | | **4 weeks** | | **8 weeks** | |
| ***Body weight and composition*** |  |  |  |  |  |  |
| Body weight, kg | 88.7 (9.22) | 94.4 (10.7) | 85.3 (9.01) | 90.4 (10.7) | 83.6 (9.20) | 87.0 (10.5) |
| Percent body fat, % | 42.4 (5.80) | 42.0 (6.76) | 41.1 (6.67) | 41.7 (6.71) | 41.1 (6.44) | 40.0 (7.55) |
| Total fat, kg | 36.1 (5.80) | 38.0 (6.53) | 33.6 (6.14) | 36 (6.22) | 33.0 (6.09) | 33.3 (6.77) |
| Arm fat, kg | 3.84 (0.77) | 4.04 (0.86) | 3.65 (0.82) | 3.93 (0.80) | 3.66 (0.75) | 3.65 (0.78) |
| Leg fat, kg | 12.1 (2.80) | 13.0 (2.92) | 11.6 (3.12) | 12.4 (2.64) | 11.3 (2.90) | 11.4 (2.44) |
| Trunk fat, kg | 19.3 (3.82) | 20.0 (4.87) | 17.4 (3.44) | 18.8 (4.72) | 17.1 (3.67) | 17.3 (5.17) |
| Android fat, kg | 3.33 (0.79) | 3.40 (0.95) | 2.93 (0.67) | 3.19 (0.99) | 2.89 (0.77) | 2.86 (1.01) |
| Gynoid fat, kg | 6.27 (1.30) | 6.82 (1.29) | 5.85 (1.42) | 6.45 (1.14) | 5.73 (1.33) | 5.83 (1.20) |
| Visceral adipose tissue, kg | 1.10 (0.58) | 1.02 (0.63) | 0.89 (0.45) | 0.93 (0.59) | 0.89 (0.48) | 0.86 (0.56) |
| Subcutaneous adipose tissue, kg | 2.54 (0.74) | 2.67 (0.72) | 2.27 (0.64) | 2.51 (0.72) | 2.23 (0.70) | 2.21 (0.77) |
| Fat free mass, kg | 52.1 (8.30) | 55.8 (9.95) | 51.2 (8.71) | 53.9 (10.0) | 50.3 (8.46) | 53.3 (10.0) |
| BMI, kg/m^2^ | 30.8 (2.20) | 31.4 (2.12) | 29.5 (1.81) | 30.2 (2.28) | 29.0 (2.06) | 29.2 (2.31) |
| ***Energy expenditure*** |  |  |  |  |  |  |
| Resting metabolic rate, kcal/d | 1944 (288) | 2016 (279) | 1843 (249) | 1972 (286) | 1843 (292) | 1915 (262) |
| RQ | 0.79 (0.03) | 0.79 (0.05) | 0.77 (0.04) | 0.76 (0.04) | 0.77 (0.04) | 0.76 (0.03) |
| Fat oxidation, g/d | 115 (34.4) | 117 (38.6) | 122 (27.2) | 141 (41.8) | 120 (37.3) | 132 (29.8) |
| Glucose oxidation, g/d | 106 (62.1) | 121 (90.9) | 65 (70.6) | 48.0 (64.9) | 68 (70.3) | 57.9 (58.2) |
| ***Urine sulfate and amino acids*** |  |  |  |  |  |  |
| Sulfate, mol/24 h | 17.6 (8.20) | 18.7 (7.48) | 29.1 (10.8) | 7.19 (3.76) | 28.9 (12.1) | 8.17 (9.32) |
| Methionine, µmol/24h | 8.98 (5.34) | 10.5 (5.75) | 9.11 (4.62) | 6.33 (3.43) | 8.33 (3.12) | 7.01 (4.84) |
| tHcy, µmol/24h | 7.99 (4.62) | 9.36 (4.99) | 8.55 (4) | 6.39 (2.99) | 7.62 (3.52) | 5.78 (2.49) |
| Cystathionine, µmol/24h | 35 (27.9) | 49 (41.5) | 37 (40.2) | 8.14 (5.26) | 39.5 (41.9) | 9.83 (10.3) |
| tCys, µmol/24h | 235 (102) | 261 (97.5) | 294 (83.2) | 257 (124) | 270 (105) | 218 (66.4) |
| ***Plasma amino acids*** |  |  |  |  |  |  |
| Methionine, µmol/L | 22.7 (3.80) | 21.3 (3.60) | 22.7 (5.18) | 20.9 (4.77) | 22.8 (2.94) | 20.4 (4.01) |
| Total homocysteine, µmol/L | 9.75 (2.62) | 9.67 (3.11) | 8.39 (2.39) | 10 (3.22) | 8.42 (2.01) | 10.4 (3.71) |
| Cystathionine, µmol/L | 0.22 (0.11) | 0.26 (0.15) | 0.29 (0.20) | 0.12 (0.05) | 0.30 (0.20) | 0.14 (0.09) |
| Total cysteine, µmol/L | 275 (43.1) | 264 (37.9) | 264 (24.9) | 267 (27.1) | 268 (33.0) | 271 (33) |
| ***Plasma biomarkers*** |  |  |  |  |  |  |
| Triglycerides, mmol/L | 1.07 (0.42) | 1.15 (0.60) | 1.07 (0.44) | 1.13 (0.43) | 1.13 (0.55) | 0.97 (0.34) |
| Total cholesterol, mmol/L | 4.93 (0.83) | 4.66 (0.75) | 4.08 (0.76) | 4.07 (0.83) | 4.31 (0.86) | 4.04 (0.81) |
| LDL, mmol/L | 3.11 (0.71) | 2.81 (0.69) | 2.55 (0.68) | 2.42 (0.69) | 2.66 (0.67) | 2.43 (0.69) |
| HDL, mmol/L | 1.39 (0.38) | 1.4 (0.36) | 1.21 (0.27) | 1.26 (0.29) | 1.23 (0.28) | 1.25 (0.26) |
| Apolipoprotein B, g/L | 0.91 (0.24) | 0.81 (0.20) | 0.76 (0.20) | 0.73 (0.21) | 0.80 (0.18) | 0.73 (0.22) |
| Apolipoprotein A1, g/L | 1.47 (0.26) | 1.49 (0.24) | 1.27 (0.17) | 1.32 (0.25) | 1.29 (0.22) | 1.29 (0.27) |
| Glucose, mmol/L | 5.31 (0.50) | 5.16 (0.41) | 5.15 (0.32) | 5.10 (0.34) | 5.11 (0.45) | 4.93 (0.32) |
| ***Plasma/serum hormones*** |  |  |  |  |  |  |
| Insulin, pmol/L | 75.5 (39.9) | 81.0 (47.0) | 55.8 (33.1) | 61.6 (33.6) | 62.0 (31.2) | 58.4 (25.1) |
| C-peptide, pmol/L | 717 (228) | 711 (214) | 611 (208) | 642 (187) | 648 (179) | 600 (175) |
| TSH, mU/L | 1.43 (0.637) | 1.6 (0.781) | 1.58 (0.748) | 1.61 (0.716) | 1.72 (0.903) | 1.54 (0.71) |
| Free T4, pmol/L | 15.9 (3.36) | 16.1 (1.87) | 16.2 (4.13) | 16.3 (1.81) | 16.4 (2.22) | 16.3 (2.09) |
| FGF-21, pg/mL | 841 (608) | 898 (1210) | 1500 (1020) | 1450 (1110) | 1420 (926) | 1290 (984) |
| IGF-1, pg/mL | 131 (56.1) | 144 (46.2) | - | - | 187 (53.5) | 185 (69.3) |
| Adiponectin, pg/mL | 18.4 (5.95) | 22.8 (9.14) | - | - | 19.9 (7.39) | 20.4 (6.28) |
| Gastric inhibitory peptide, pg/mL | 197 (248) | 181 (152) | 204 (234) | 176 (140) | 196 (235) | 209 (240) |
| Glucagon-like peptide 1, pg/mL | 71.2 (30.2) | 63.1 (26.5) | 69.5 (31.7) | 54.8 (17.7) | 66.8 (20.9) | 56.4 (15.2) |
| Ghrelin, pg/mL | 144 (87.6) | 149 (84.5) | 236 (226) | 190 (145) | 188 (114) | 169 (101) |
| Leptin, ng/mL | 51.4 (25) | 50.6 (22.8) | 39.3 (25.8) | 35.3 (19.6) | 43.2 (25.9) | 31.9 (22.2) |
| Pancreatic peptide, pg/mL | 169 (137) | 161 (150) | 148 (84.8) | 186 (191) | 160 (107) | 207 (161) |
| Peptide YY, pg/mL | 70 (44.4) | 59 (31.6) | 79.1 (45) | 61.8 (31.2) | 75.3 (40.7) | 62.9 (37) |
| ***Ketone bodies*** |  |  |  |  |  |  |
| β-hydroxybutyrate, µmol/L | 35.8 (47.6) | 69.9 (114) | 82 (55.7) | 151 (155) | 80.9 (102) | 214 (384) |
| Acetoacetate, µmol/L | 18.4 (14.8) | 25.2 (32.6) | 33 (18.1) | 51.8 (47) | 35.1 (32.2) | 68 (87.2) |
| Acetone, µmol/L | 13.7 (3.67) | 16.5 (11.9) | 22.3 (10.8) | 27.6 (23.1) | 21.5 (14.3) | 31.8 (30.2) |
| Acetate, µmol/L | 40.8 (21.4) | 45.8 (23.4) | 50 (26.1) | 60.3 (28.7) | 52.3 (26.0) | 56.2 (22.8) |
| ***Safety markers*** |  |  |  |  |  |  |
| ASAT, U/L | 22.6 (6.93) | 24.3 (8.83) | 21.8 (6.21) | 25.5 (12) | 21.5 (5.32) | 23 (5.15) |
| ALAT, U/L | 27.9 (17.3) | 25.4 (9.64) | 28.7 (19.7) | 28.3 (21.2) | 23.9 (11.0) | 22.5 (7.46) |
| Urea, mmol/L | 4.31 (1.26) | 4.26 (1.00) | 3.2 (1.01) | 2.81 (0.68) | 3.24 (1.00) | 3.17 (0.96) |
| Creatinine, µmol/L | 69.0 (11.9) | 66.4 (11.4) | 66.1 (10.8) | 67.4 (10.8) | 63.8 (11.7) | 66.5 (8.40) |
| Vitamin B12, pmol/L | 398 (193) | 367 (133) | 380 (178) | 402 (120) | 429 (143) | 413 (125) |
| Methylmalonic acid, µmol/L | 0.19 (0.07) | 0.18 (0.06) | 0.16 (0.04) | 0.16 (0.05) | 0.15 (0.04) | 0.19 (0.13) |
| ^1^ Abbreviations: ALAT, alanine aminotransferase; ASAT, aspartate aminotransferase; BMI, body mass index; FGF-21, fibroblast growth factor 21; IGF-1, insulin-like growth factor 1; RMR, resting metabolic rate expenditure; RQ, respiratory quotient; tCys, total cysteine; tHcy, total homocysteine | | | | | | |

| Supplemental Table S4. Median (25^th^, 75^th^ %-tile) observed values for all outcomes in the SAAR and control groups^1^. | | | | | | |
| --- | --- | --- | --- | --- | --- | --- |
|  | **Control** | **SAAR** | **Control** | **SAAR** | **Control** | **SAAR** |
|  | **Baseline** | | **4 weeks** | | **8 weeks** | |
| ***Body weight and composition*** |  |  |  |  |  |  |
| Body weight, kg | 88.6 (82.0, 95.7) | 93.2 (86.6, 102) | 86.4 (78.0, 92.1) | 88.8 (82.4, 98.2) | 84.0 (77.7, 89.6) | 86 (80.2, 91.0) |
| Percent body fat, % | 44.1 (39.2, 46.4) | 44.0 (37.8, 47.3) | 41.8 (36.9, 46.3) | 42.2 (37.1, 47.0) | 42.7 (37.1, 45.6) | 41.0 (35.2, 45.5) |
| Total fat, kg | 37.2 (31.0, 41.5) | 37.9 (34.4, 42.8) | 33.7 (29.0, 39.4) | 35.5 (31.8, 40.2) | 34.5 (28.5, 38.0) | 33.9 (29.7, 36.6) |
| Arm fat, kg | 3.82 (3.42, 4.20) | 4.04 (3.46, 4.73) | 3.62 (3.25, 4.04) | 3.64 (3.36, 4.46) | 3.61 (3.22, 4.23) | 3.53 (3.14, 4.39) |
| Leg fat, kg | 12.0 (10.8, 13.9) | 12.7 (11.4, 14.5) | 12.0 (10.0, 13.9) | 11.9 (10.8, 13.5) | 11.1 (9.77, 12.9) | 11.0 (10.1, 12.5) |
| Trunk fat, kg | 19.8 (15.8, 22.2) | 20.2 (16.7, 22.5) | 17.3 (14.4, 20.6) | 18.5 (16.1, 21.0) | 17.0 (14.4, 20.0) | 16.8 (14.0, 19.2) |
| Android fat, kg | 3.43 (2.71, 4.00) | 3.45 (2.80, 3.79) | 3.02 (2.43, 3.4) | 3.25 (2.54, 3.51) | 3.00 (2.25, 3.42) | 2.83 (2.26, 3.29) |
| Gynoid fat, kg | 6.29 (5.4, 7.04) | 6.83 (5.95, 7.49) | 5.83 (4.92, 6.78) | 6.53 (5.48, 7.28) | 5.70 (4.85, 6.48) | 6.07 (4.83, 6.67) |
| Visceral adipose tissue, kg | 1.12 (0.77, 1.32) | 0.88 (0.58, 1.16) | 0.97 (0.69, 1.08) | 0.83 (0.63, 1.03) | 0.86 (0.60, 1.11) | 0.77 (0.52, 1.04) |
| Subcutaneous adipose tissue, kg | 2.49 (1.93, 3.11) | 2.73 (2.22, 3.08) | 2.13 (1.75, 2.87) | 2.54 (1.98, 2.92) | 2.23 (1.66, 2.83) | 2.11 (1.77, 2.54) |
| Fat free mass, kg | 48.8 (46.7, 55.6) | 53.8 (49.0, 62.2) | 50.0 (45.6, 54.3) | 51.2 (47.0, 59.4) | 47.3 (45.3, 52.9) | 51.3 (46.6, 59.0) |
| BMI, kg/m^2^ | 30.3 (29, 32.4) | 31.2 (29.5, 33.2) | 29.0 (28.2, 30.8) | 29.6 (28.5, 32.2) | 28.7 (27.1, 30.7) | 28.8 (27.2, 31.2) |
| ***Energy expenditure*** |  |  |  |  |  |  |
| Resting metabolic rate, kcal/d | 1944 (1728, 2145) | 2016 (1814, 2131) | 1857 (1612, 2016) | 1886 (1786, 2117) | 1814 (1656, 2074) | 1872 (1785, 2073) |
| Fat oxidation, g/d | 110 (93.6, 135) | 118 (91.9, 145) | 114 (108, 131) | 137 (109, 170) | 123 (97.9, 132) | 132 (115, 148) |
| Glucose oxidation, g/d | 118 (59.6, 154) | 93.3 (74.5, 171) | 48.5 (23.6, 111) | 53.0 (10.1, 74.7) | 66.0 (4.44, 123) | 53.9 (27.8, 107) |
| Respiratory quotient | 0.786 (0.771, 0.807) | 0.788 (0.763, 0.825) | 0.754 (0.745, 0.789) | 0.754 (0.736, 0.779) | 0.77 (0.737, 0.796) | 0.758 (0.747, 0.781) |
| ***Urine sulfur amino acids*** |  |  |  |  |  |  |
| Sulfate mmol/24 h | 15.2 (12.4, 22.1) | 19.2 (13.7, 21.4) | 31.5 (21.4, 34.9) | 5.6 (4.61, 9.09) | 29.2 (21.2, 34.5) | 6.43 (4.95, 8.05) |
| Methionine, µmol/24 h | 7.37 (5.51, 12.0) | 8.98 (6.44, 14.1) | 8.94 (5.36, 11.6) | 5.45 (3.8, 8.12) | 7.85 (6.95, 10.40) | 5.76 (4.04, 8.56) |
| tHcy, µmol/24 h | 6.75 (4.57, 9.95) | 7.96 (6.28, 11.3) | 7.22 (6.25, 10.4) | 5.82 (4.17, 6.99) | 6.76 (5.43, 9.27) | 5.07 (4.42, 6.93) |
| Cystathionine, µmol/24 h | 24.4 (18.9, 40.5) | 33.5 (19.4, 63.6) | 20.1 (14.7, 44.7) | 7.46 (4.13, 10.8) | 24 (15.3, 43.7) | 5.93 (3.05, 10.4) |
| tCys, µmol/24 h | 233 (186, 266) | 241 (206, 310) | 302 (246, 344) | 217 (184, 306) | 247 (205, 324) | 223 (178, 253) |
| ***Plasma amino acids*** |  |  |  |  |  |  |
| Methionine, µmol/L | 22.7 (20.1, 26.0) | 20.7 (19.3, 23.2) | 22.2 (18.7, 25.3) | 19.3 (17.9, 23.9) | 22.7 (21.0, 24.2) | 19.8 (17.7, 22.2) |
| Total homoycsteine, µmol/L | 9.33 (7.90, 11.1) | 9.34 (7.42, 10.8) | 6.94 (6.68, 10.7) | 9.39 (8.03, 11.1) | 8.13 (6.98, 9.67) | 9.51 (8.54, 11.6) |
| Cystathionine, µmol/L | 0.19 (0.16, 0.27) | 0.22 (0.16, 0.32) | 0.19 (0.15, 0.39) | 0.12 (0.08, 0.15) | 0.22 (0.17, 0.34) | 0.11 (0.09, 0.14) |
| Total cysteine, µmol/L | 269 (244, 290) | 261 (242, 273) | 267 (245, 284) | 267 (246, 284) | 263 (253, 276) | 271 (255, 283) |
| ***Plama biomarkers*** |  |  |  |  |  |  |
| Triglycerides, mmol/L | 1.00 (0.80, 1.35) | 1.00 (0.70, 1.30) | 0.95 (0.80, 1.30) | 1.00 (0.90, 1.40) | 0.90 (0.78, 1.22) | 0.90 (0.80, 1.17) |
| Total cholesterol, mmol/L | 5.00 (4.35, 5.35) | 4.90 (4.00, 5.15) | 4.05 (3.52, 4.68) | 4.10 (3.50, 4.65) | 4.20 (3.68, 5.05) | 3.95 (3.52, 4.38) |
| LDL, mmol/L | 3.20 (2.65, 3.50) | 2.80 (2.15, 3.35) | 2.45 (2.10, 3.08) | 2.20 (1.85, 2.95) | 2.55 (2.10, 3.20) | 2.20 (1.92, 2.77) |
| HDL, mmol/L | 1.30 (1.15, 1.50) | 1.30 (1.10, 1.65) | 1.20 (1.10, 1.37) | 1.30 (1.00, 1.45) | 1.20 (1.00, 1.42) | 1.15 (1.1, 1.48) |
| ApolipoproteinB, g/L | 0.90 (0.80, 1.00) | 0.80 (0.65, 0.90) | 0.70 (0.60, 0.90) | 0.70 (0.60, 0.80) | 0.75 (0.70, 0.90) | 0.70 (0.60, 0.80) |
| Apolipoprotein A1, g/L | 1.40 (1.30, 1.55) | 1.50 (1.30, 1.60) | 1.25 (1.20, 1.37) | 1.30 (1.10, 1.45) | 1.30 (1.10, 1.40) | 1.20 (1.10, 1.40) |
| ***Plasma hormones*** |  |  |  |  |  |  |
| Glucose, mmol/L | 5.40 (5.05, 5.60) | 5.20 (4.90, 5.45) | 5.15 (5.03, 5.40) | 5.10 (4.80, 5.40) | 5.05 (4.80, 5.40) | 5.00 (4.70, 5.18) |
| Insulin, pmol/L | 66.0 (47.5, 99.5) | 77.0 (43.5, 96.0) | 47.5 (36.5, 66.5) | 49.0 (36.5, 82.5) | 54.0 (42.8, 77.8) | 54.5 (40.2, 71.2) |
| C-peptide, pmol/L | 665 (612, 843) | 688 (574, 857) | 628 (517, 682) | 650 (548, 740) | 575 (539, 722) | 616 (480, 720) |
| Free T4, pmol/L | 16.0 (15.0, 18.0) | 16.0 (15.0, 17.0) | 17.0 (15.2, 18.0) | 16.0 (15.0, 17.0) | 16.0 (15.0, 17.2) | 16.0 (14.2, 18.0) |
| TSH, mU/L | 1.30 (1.10, 1.85) | 1.40 (1.00, 2.20) | 1.55 (1.05, 2.12) | 1.30 (1.05, 2.25) | 1.40 (1.20, 2.08) | 1.35 (1.10, 1.67) |
| FGF-21, pg/mL | 631 (452, 1040) | 597 (259, 888) | 1120 (866, 1900) | 1050 (642, 2060) | 1010 (846, 1850) | 908 (493, 2010) |
| IGF-1, pg/mL | 127 (81.5, 157) | 140 (123, 175) | - | - | 177 (150, 217) | 198 (137, 234) |
| Adiponectin, pg/mL | 17.5 (14.4, 22.0) | 23.6 (14.9, 28.7) | - | - | 17.6 (14.9, 23.3) | 20.6 (16.2, 24.8) |
| Gastric inhibitory peptide, pg/mL | 129 (112, 209) | 140 (103, 195) | 147 (121, 181) | 137 (107, 200) | 136 (118, 183) | 136 (107, 183) |
| Glucagon-like peptide 1, pg/mL | 63.7 (50.5, 94.5) | 58.8 (45.1, 74.4) | 58.7 (49.7, 83.2) | 51.3 (42.7, 60.7) | 61.6 (53.3, 78) | 54.6 (42.9, 69.0) |
| Ghrelin, pg/mL | 122 (81.1, 173) | 118 (90.4, 189) | 159 (134, 239) | 141 (104, 252) | 173 (106, 248) | 134 (98.4, 202) |
| Leptin, ng/mL | 54.9 (26.2, 71.3) | 50.5 (35.1, 69.6) | 45.0 (13.4, 61.4) | 37.2 (23.0, 50.4) | 44.7 (17.2, 63.2) | 27.0 (12.5, 45.0) |
| Pancreatic peptide, pg/mL | 137 (72.7, 169) | 106 (88.3, 156) | 130 (89.4, 185) | 124 (89.6, 185) | 136 (80.7, 228) | 172 (101, 231) |
| Peptide YY, pg/mL | 62.2 (43.4, 77.6) | 59.3 (36.7, 73.9) | 58.7 (51.8, 92.9) | 54.8 (47.2, 71.6) | 64.3 (58.8, 88.8) | 49.2 (39.9, 83.2) |
| ***Ketone bodies*** |  |  |  |  |  |  |
| β-hydroxybutyrate, µmol/L | 22.0 (9.41, 32.6) | 23.9 (6.20, 80.1) | 68.1 (40.6, 129) | 100 (32.3, 235) | 42 (33.9, 90.8) | 117 (68.3, 229) |
| Acetoacetate, µmol/L | 14.9 (9, 23.1) | 13.4 (8.71, 26.2) | 28.2 (18.1, 46.9) | 31.4 (15.8, 77.3) | 23.8 (13.7, 49.7) | 48.9 (27.4, 73.9) |
| Acetone, µmol/L | 12.6 (11.8, 14.8) | 14.0 (11.9, 15.7) | 18.5 (16.1, 25.6) | 19.8 (15.3, 30.1) | 18.0 (13.4, 24.3) | 24.8 (17.4, 36.0) |
| Acetate, µmol/L | 37.3 (24.3, 44.6) | 42.6 (26.7, 61.2) | 39.6 (30.8, 59.5) | 49.9 (36.4, 82.6) | 45.3 (35.1, 65.9) | 53.5 (37.8, 70.9) |
| ***Safety markers*** |  |  |  |  |  |  |
| ASAT, U/L | 20.0 (18.0, 25.5) | 24.0 (19.0, 26.0) | 19.5 (17.2, 24.0) | 23.0 (20.0, 27.8) | 22.0 (18.0, 25.0) | 21.5 (20.0, 24.8) |
| ALAT, U/L | 22.0 (17.5, 31.0) | 24.0 (20.0, 27.0) | 19.0 (17.2, 32.5) | 21.0 (18.0, 32,0) | 20.0 (16.0, 26.0) | 22.5 (17.2, 24.8) |
| Urea, mmol/L | 4.20 (3.35, 4.70) | 4.4 (3.60, 4.85) | 2.90 (2.62, 3.4.0) | 2.80 (2.45, 3.10) | 2.85 (2.50, 3.65) | 3.15 (2.6.0, 3.58) |
| Creatinine, µmol/L | 72.0 (60.5, 76.5) | 67 (56.5, 74.5) | 65.0 (60.0, 75.0) | 68,0 (60.5, 75.5) | 63 (52.8, 71.5) | 66.5 (62.2, 70.5) |
| Vitamin B12, pmol/L | 375 (293, 450) | 328 (283, 444) | 386 (273, 448) | 377 (316, 456) | 394 (323, 486) | 410 (320, 478) |
| Methylmalonic acid, µmol/L | 0.18 (0.14, 0.21) | 0.17 (0.13, 0.21) | 0.15 (0.12, 0.19) | 0.14 (0.12, 0.19) | 0.14 (0.12, 0.165) | 0.16 (0.13, 0.19) |
| ^1^ Abbreviations: ALAT, alanine aminotransferase; ASAT, aspartate aminotransferase; BMI, body mass index; FGF-21, fibroblast growth factor 21; IGF-1, insulin-like growth factor 1; RMR, resting metabolic rate expenditure; RQ, respiratory quotient; tCys, total cysteine; tHcy, total homocysteine | | | | | | |

| Supplemental Table S5. Estimated marginal means (95 % CI) body composition in the SAAR and control groups^1^. | | | | | | |
| --- | --- | --- | --- | --- | --- | --- |
|  | **Group** | **Baseline** | **4 weeks** | **8 weeks** | **p_int 4 weeks_** | **p_int_ _8 weeks_** |
| ***Body weight and composition*** |  |  |  |  |  |  |
| Body weight, kg | Control | 94.1  (91.5, 96.7) | 90.7  (88.0, 93.3) | 88.7  (86.1, 91.4) | 0.13 | 0.012 |
|  | SAAR | 94.1  (91.5, 96.7) | 90.0  (87.3, 92.6) | 87.6  (84.9, 90.2) |  |  |
| BMI, kg/m^2^ | Control | 30.9  (30.3, 31.5) | 29.7  (29.1, 30.4) | 29.1  (28.4, 29.7) | 0.23 | 0.049 |
|  | SAAR | 30.9  (30.3, 31.5) | 29.6  (28.9, 30.2) | 28.8  (28.1, 29.4) |  |  |
| Body fat, % | Control | 39.5  (38.3, 40.8) | 38.9  (37.7, 40.2) | 38.0  (36.8, 39.3) | 0.79 | 0.98 |
|  | SAAR | 39.5  (38.3, 40.8) | 38.9  (37.6, 40.1) | 38.0  (36.8, 39.3) |  |  |
| Total fat, kg | Control | 35.7  (34.0, 37.4) | 33.8  (32.0, 35.5) | 32.3  (30.6, 34.1) | 0.41 | 0.27 |
|  | SAAR | 35.7  (34.0, 37.4) | 33.5  (31.7, 35.2) | 31.9  (30.2, 33.7) |  |  |
| Arm fat, kg | Control | 3.74  (3.53, 3.95) | 3.66  (3.44, 3.88) | 3.51  (3.29, 3.73) | 0.67 | 0.68 |
|  | SAAR | 3.74  (3.53, 3.95) | 3.64  (3.42, 3.85) | 3.49  (3.27, 3.7) |  |  |
| Leg fat, kg | Control | 11.7  (11.0, 12.3) | 11.2  (10.6, 11.9) | 10.8  (10.1, 11.4) | 0.50 | 0.31 |
|  | SAAR | 11.7  (11.0, 12.3) | 11.1  (10.5, 11.8) | 10.6  (9.92, 11.3) |  |  |
| Trunk fat, kg | Control | 19.4  (18.1, 20.6) | 17.9  (16.6, 19.2) | 17.1  (15.8, 18.4) | 0.62 | 0.46 |
|  | SAAR | 19.4  (18.1, 20.6) | 17.8  (16.5, 19.1) | 16.9  (15.6, 18.2) |  |  |
| Android fat, kg | Control | 3.35  (3.1, 3.61) | 3.05  (2.79, 3.31) | 2.91  (2.65, 3.17) | 0.80 | 0.38 |
|  | SAAR | 3.35  (3.1, 3.61) | 3.07  (2.81, 3.33) | 2.86  (2.6, 3.12) |  |  |
| Gynoid fat, kg | Control | 6.16  (5.85, 6.48) | 5.80  (5.48, 6.13) | 5.57  (5.24, 5.89) | 0.66 | 0.060 |
|  | SAAR | 6.16  (5.85, 6.48) | 5.77  (5.45, 6.1) | 5.44  (5.12, 5.76) |  |  |
| Visceral adipose tissue, kg | Control | 1.17  (1.03, 1.32) | 0.98  (0.83, 1.14) | 0.95  (0.80, 1.11) | 0.21 | 0.57 |
|  | SAAR | 1.17  (1.03, 1.32) | 1.04  (0.89, 1.19) | 0.99  (0.83, 1.13) |  |  |
| Subcutaneous adipose tissue, kg | Control | 2.50  (2.30, 2.70) | 2.33  (2.12, 2.54) | 2.19  (1.98, 2.4) | 0.55 | 0.12 |
|  | SAAR | 2.50  (2.30, 2.70) | 2.30  (2.09, 2.51) | 2.11 (1.91, 2.32) |  |  |
| Fat free mass, kg | Control | 57.9  (56.3, 59.4) | 56.4  (54.9, 58) | 56.1  (54.5, 57.7) | 0.13 | 0.013 |
|  | SAAR | 57.9  (56.3, 59.4) | 56.1  (54.5, 57.6) | 55.4  (53.9, 57.0) |  |  |
| ***Energy expenditure*** |  |  |  |  |  |  |
| RMR, kcal/day^2^ | Control | 1976  (1917, 2035) | 1911  (1847, 1976) | 1914  (1851, 1977) | 0.14 | 0.69 |
|  | SAAR | 1976  (1917, 2035) | 1956  (1891, 2021) | 1926  (1862, 1991) |  |  |
| Fat oxidation, g/day | Control | 122 (4.69) | 125 (7.21) | 124 (7.00) | 0.031 | 0.16 |
|  | SAAR | 122 (4.69) | 145 (6.63) | 137 (6.73) |  |  |
| Carbohydrate oxidation, g/day | Control | 126 (9.35) | 80.2 (14.5) | 86.2 (14.1) | 0.25 | 0.33 |
|  | SAAR | 126 (9.35) | 58.8 (13.3) | 68.3 (13.5) |  |  |
| Respiratory quotient | Control | 0.79  (0.78, 0.80) | 0.77  (0.75, 0.78) | 0.77  (0.76, 0.79) | 0.14 | 0.69 |
|  | SAAR | 0.79  (0.78, 0.80) | 0.76  (0.74, 0.77) | 0.76  (0.75, 0.78) |  |  |
| ***Plasma sulfur amino acids, µmol/L*** |  |  |  |  |  |  |
| Methionine | Control | 22.6  (21.5, 23.7) | 22.5  (21.0, 23.9) | 22.7  (21.3, 24.1) | 0.49 | 0.14 |
|  | SAAR | 22.6  (21.5, 23.7) | 21.9  (20.5, 23.3) | 21.4  (20.0, 22.9) |  |  |
| Total homocysteine | Control | 10.1  (9.32, 10.9) | 8.64  (7.71, 9.56) | 8.84  (7.92, 9.75) | < 0.001 | < 0.001 |
|  | SAAR | 10.1  (9.32, 10.9) | 10.5  (9.61, 11.4) | 10.9  (10.0, 11.8) |  |  |
| Cystathionine | Control | 0.25  (0.21, 0.29) | 0.29  (0.24, 0.35) | 0.31  (0.26, 0.37) | < 0.001 | < 0.001 |
|  | SAAR | 0.25  (0.214, 0.29) | 0.13  (0.08, 0.18) | 0.15  (0.10, 0.20) |  |  |
| Total cysteine | Control | 272  (262, 281) | 261  (248, 274) | 266  (254, 279) | 0.09 | 0.11 |
|  | SAAR | 272  (262, 281) | 274  (262, 286) | 279  (266, 291) |  |  |
| ***Sulfur amino acid output*** |  |  |  |  |  |  |
| Methionine, µmol/24h | Control | 10.5  (9.29, 11.8) | 10.2  (8.45, 11.9) | 9.92  (8.17, 11.7) | < 0.001 | 0.014 |
|  | SAAR | 10.5  (9.29, 11.8) | 6.33  (4.69, 7.97) | 7.22  (5.55, 8.88) |  |  |
| Total homocysteine, µmol/24h | Control | 9.42  (8.38, 10.5) | 9.51  (8.07, 10.9) | 9.24  (7.78, 10.7) | < 0.001 | < 0.001 |
|  | SAAR | 9.42  (8.38, 10.5) | 6.47  (5.10, 7.84) | 6.21  (4.81, 7.60) |  |  |
| Cystathionine, µmol/24h | Control | 44.5  (36.0, 53.0) | 39.3  (26.8, 51.9) | 43.7  (30.9, 56.5) | < 0.001 | < 0.001 |
|  | SAAR | 44.5  (36.0, 53.0) | 8.96  (-2.90, 20.8) | 11.5  (-0.61, 23.6) |  |  |
| Total cysteine, µmol/24h | Control | 265  (238, 291) | 313  (275, 351) | 307  (268, 345) | 0.03 | 0.002 |
|  | SAAR | 265  (238, 291) | 260  (225, 296) | 229  (193, 266) |  |  |
| Sulfate, mol/24 h | Control | 18.8  (16.3, 21.3) | 28.7  (25.1, 32.3) | 29.6  (25.8, 33.5) | < 0.001 | < 0.001 |
|  | SAAR | 18.8  (16.3, 21.3) | 7.71  (4.27, 11.2) | 8.88  (5.35, 12.4) |  |  |
| ***Plasma biomarkers*** |  |  |  |  |  |  |
| Triglycerides, mmol/L | Control | 1.13  (0.99, 1.27) | 1.18  (1.00, 1.35) | 1.19  (1.02, 1.36) | 0.90 | 0.15 |
|  | SAAR | 1.13  (0.99, 1.27) | 1.19  (1.02, 1.36) | 1.06  (0.89, 1.23) |  |  |
| Total cholesterol, mmol/L | Control | 4.79  (4.55,.02) | 4.12  (3.83, 4.41) | 4.24  (3.95, 4.52) | 0.75 | 0.67 |
|  | SAAR | 4.79  (4.55, 5.02) | 4.17  (3.89, 4.45) | 4.17  (3.89, 4.45) |  |  |
| LDL, mmol/L | Control | 3.00  (2.80, 3.20) | 2.59  (2.34, 2.83) | 2.60  (2.37, 2.84) | 0.90 | 0.70 |
|  | SAAR | 3.00  (2.80, 3.20) | 2.57  (2.34, 2.80) | 2.56  (2.32, 2.79) |  |  |
| HDL, mmol/L | Control | 1.35  (1.27, 1.44) | 1.17  (1.06, 1.27) | 1.19  (1.09, 1.29) | 0.35 | 0.29 |
|  | SAAR | 1.35  (1.27, 1.44) | 1.21  (1.11, 1.31) | 1.24  (1.14, 1.34) |  |  |
| Apolipoprotein B, g/L | Control | 0.88  (0.81, 0.94) | 0.78  (0.71, 0.86) | 0.80  (0.72, 0.87) | 0.78 | 0.76 |
|  | SAAR | 0.88  (0.81, 0.94) | 0.79  (0.72, 0.87) | 0.78  (0.71, 0.86) |  |  |
| Apolipoprotein A1, g/l | Control | 1.44  (1.37, 1.51) | 1.26  (1.18, 1.34) | 1.3  (1.21, 1.38) | 0.52 | 0.82 |
|  | SAAR | 1.44  (1.37, 1.51) | 1.29  (1.21, 1.37) | 1.29  (1.21, 1.37) |  |  |
| Fasting glucose, mmol/L | Control | 5.28  (5.17, 5.39) | 5.16  (5.01, 5.32) | 5.10  (4.95, 5.24) | 0.98 | 0.42 |
|  | SAAR | 5.28  (5.17, 5.39) | 5.16  (5.02, 5.30) | 5.02  (4.88, 5.17) |  |  |
| ***Plasma hormones*** |  |  |  |  |  |  |
| Fasting insulin, pmol/L | Control | 75.4  (65.4, 85.4) | 58.2  (44.3, 72.1) | 61.4  (47.8, 74.9) | 0.80 | 0.28 |
|  | SAAR | 75.4  (65.4, 85.4) | 56  (42.9, 69.1) | 52.1  (38.8, 65.5) |  |  |
| C-peptide, pmol/L | Control | 697  (642, 752) | 613  (535, 691) | 638  (563, 714) | 0.98 | 0.22 |
|  | SAAR | 697  (642, 752) | 615  (541, 688) | 579  (505, 653) |  |  |
| TSH, mU/L | Control | 1.50  (1.29, 1.70) | 1.62  (1.34, 1.9) | 1.79  (1.52, 2.07) | 0.70 | 0.08 |
|  | SAAR | 1.50  (1.29, 1.70) | 1.56  (1.29, 1.82) | 1.49  (1.22, 1.76) |  |  |
| Free T4, pmol/L | Control | 16.4  (15.7, 17.1) | 16.8  (15.8, 17.8) | 16.9  (16, 17.9) | 0.90 | 0.64 |
|  | SAAR | 16.4 (15.7, 17.1) | 16.9  (15.9, 17.8) | 16.6  (15.7, 17.6) |  |  |
| FGF-21, pg/mL | Control | 596  (482, 735) | 1140  (886, 1470) | 1030  (800, 1320) | 0.91 | 0.63 |
|  | SAAR | 596  (482, 735) | 1160  (908, 1480) | 1100  (856, 1400) |  |  |
| IGF-1, pg/mL | Control | 141  (124, 157) | - | 194  (170, 217) | - | 0.46 |
|  | SAAR | 141  (124, 157) | - | 183  (160, 205) |  |  |
| Adiponectin, pg/mL | Control | 19.9  (17.7, 22.1) | - | 20.2  (17.5, 22.9) | - | 0.35 |
|  | SAAR | 19.9  (17.7, 22.1) | - | 18.8  (16.1, 21.4) |  |  |
| Gastric inhibitory peptide, pg/mL | Control | 183  (124, 242) | 185  (118, 252) | 184  (118, 251) | 0.69 | 0.41 |
|  | SAAR | 183  (124, 242) | 173  (108, 239) | 208  (143, 274) |  |  |
| Glucagon-like peptide 1, pg/mL | Control | 65.2  (58.2, 72.2) | 66.5  (57.7, 75.4) | 64.4  (55.7, 73.1) | 0.017 | 0.11 |
|  | SAAR | 65.2  (58.2, 72.2) | 54.8  (46.4, 63.3) | 56.5  (48.0, 65.1) |  |  |
| Leptin, ng/mL | Control | 43.6  (38.3, 49) | 32.9  (26.8, 39) | 32.0  (25.9, 38.1) | 0.14 | 0.03 |
|  | SAAR | 43.6  (38.3, 49) | 28.8  (22.9, 34.8) | 26.0  (20.0, 32.0) |  |  |
| Pancreatic peptide, pg/mL | Control | 162  (121, 202) | 143  (87.7, 198) | 152  (97.8, 206) | 0.24 | 0.16 |
|  | SAAR | 162  (121, 202) | 182  (130, 234) | 199  (147, 252) |  |  |
| Peptide YY, pg/mL | Control | 62.1  (51.2, 73) | 70.0  (56.9, 83) | 65.3  (52.4, 78.1) | 0.37 | 0.91 |
|  | SAAR | 62.1  (51.2, 73) | 64.1  (51.5, 76.6) | 64.6  (51.9, 77.2) |  |  |
| Ghrelin, pg/mL | Control | 146  (101, 192) | 201  (151, 252) | 187  (136, 237) | 0.76 | 0.56 |
|  | SAAR | 146  (101, 192) | 195  (145, 244) | 175  (125, 225) |  |  |
| ***Ketone bodies*** |  |  |  |  |  |  |
| β-hydroxybutyrate, µmol/L | Control | 22.9  (16.3, 32.2) | 66.9  (40.5, 110) | 54.5  (33.3, 89.1) | 0.37 | 0.023 |
|  | SAAR | 22.9  (16.3, 32.2) | 88.4  (55.1, 142) | 114  (70.9, 183) |  |  |
| Acetoacetate, µmol/L | Control | 14.7  (11.8, 18.3) | 31.4  (22.5, 43.8) | 29.1  (21, 40.4) | 0.40 | 0.042 |
|  | SAAR | 14.7  (11.8, 18.3) | 35.0  (25.6, 47.9) | 45.4  (33.2, 62.2) |  |  |
| Acetone, µmol/L | Control | 17.2  (12.2, 22.1) | 25.6  (18.2, 33.0) | 24.8  (17.5, 32.0) | 0.39 | 0.085 |
|  | SAAR | 17.2  (12.2, 22.1 | 29.9  (22.9, 36.9) | 33.2  (26.2, 40.2) |  |  |
| Acetate, µmol/L | Control | 43.5  (46.5, 50.6) | 52.8  (42.4, 63.1) | 53.6  (43.5, 63.8) | 0.36 | 0.59 |
|  | SAAR | 43.5  (46.5, 50.6) | 59.1  (49.3, 69.0) | 57.3  (47.5, 67.1) |  |  |
| ***Safety markers*** |  |  |  |  |  |  |
| ASAT, U/L | Control | 24.4  (22.3, 26.5) | 23.0  (19.9, 26.2) | 23.1  (20.0, 26.1) | 0.13 | 0.71 |
|  | SAAR | 24.4  (22.3, 26.5) | 26.2  (23.3, 29.2) | 23.8  (20.9, 26.8) |  |  |
| ALAT, U/L | Control | 29.1  (25.1, 33.1) | 30.0  (24.5, 35.5) | 27.6  (22.2, 32.9) | 0.67 | 0.57 |
|  | SAAR | 29.1  (25.1, 33.1) | 31.4  (26.2, 36.6) | 25.7  (20.4, 31.0) |  |  |
| Urea, mmol/L | Control | 4.44  (4.17, 4.71) | 3.33  (2.99, 3.67) | 3.40  (3.07, 3.73) | 0.14 | 0.59 |
|  | SAAR | 4.44  (4.17, 4.71) | 3.06  (2.74, 3.38) | 3.31  (2.98, 3.63) |  |  |
| Creatinine, µmol/L | Control | 70.4  (67.8, 73.1) | 68.2  (65.1, 71.2) | 66.7  (63.7, 69.7) | 0.01 | 0.01 |
|  | SAAR | 70.4  (67.8, 73.1) | 72.0  (69.0, 74.9) | 70.3  (67.4, 73.3) |  |  |
| Vitamin B12, pmol/L | Control | 380  (338, 423) | 364  (311, 417) | 401  (349, 453) | 0.06 | 0.75 |
|  | SAAR | 380  (338, 423) | 419  (368, 469) | 410  (359, 462) |  |  |
| MMA, µmol/L | Control | 0.18  (0.16, 0.20) | 0.15  (0.13, 0.18) | 0.16  (0.13, 0.18) | 0.85 | 0.17 |
|  | SAAR | 0.18  (0.16, 0.20) | 0.16  (0.13, 0.19) | 0.18  (0.15, 0.21) |  |  |
| ^1^ Estimated means and p-values are derived from linear mixed models with body mass as the outcome, and group, visit and their interaction term (group × time) at 4 and 8 weeks as predictors. The models were baseline adjusted. The p-value indicates the difference between groups at each timepoint. Abbreviations: BMI, body mass index; SAAR, sulfur amino acid restriction  ^2^ Additionally adjusted for total fat mass | | | | | | |

#### 2.3. Per-protocol analyses

| Supplemental Table S6. Baseline characteristics of completers in the SAAR and control groups^1^. | | | | |
| --- | --- | --- | --- | --- |
|  | **SAAR**  **n = 28** | | **Control**  **n = 26** | |
| Males, n (%) | 7 (25.0) | | 7 (26.9) | |
|  | **Mean (SD)** | **Median (IQR)** | **Mean (SD)** | **Median (IQR)** |
| Age, y | 32.7 (5.80) | 31 (29, 36) | 34.8 (6.32) | 34.5 (30.2, 40.2) |
| Body weight, kg | 94.1 (11) | 93.1 (86, 101) | 88.9 (9.50) | 89.7 (80.6, 95.9) |
| Body mass index, kg/m^2^ | 31.4 (2.19) | 31.2 (29.5, 33.3) | 30.7 (2.20) | 30.3 (28.9, 32.3) |
| Waist-to-hip ratio | 0.86 (0.08) | 0.84 (0.80, 0.92) | 0.86 (0.08) | 0.84 (0.90, 0.91) |
| Percent body fat, % | 42.0 (6.95) | 43.9 (38.4, 47.4) | 42.6 (5.57) | 44.1 (39.4, 46.4) |
| Systolic blood pressure, mm/Hg | 120 (11.4) | 117 (111, 127) | 119 (10.7) | 118 (111, 125) |
| Diastolic blood pressure, mm/Hg | 69.2 (7.89) | 69.8 (64.3, 74) | 69.8 (8.16) | 66.7 (63.8, 75.7) |
| Apolipoprotein B, g/L | 0.80 (0.20) | 0.80 (0.60, 0.90) | 0.92 (0.23) | 0.90 (0.80, 1.00) |
| Apolipoprotein A1, g/L | 1.48 (0.25) | 1.40 (1.30, 1.60) | 1.48 (0.26) | 1.40 (1.30, 1.60) |
| LDL, mmol/L | 2.79 (0.704) | 2.85 (2.10, 3.32) | 3.14 (0.686) | 3.20 (2.70, 3.50) |
| HDL, mmol/L | 1.42 (0.375) | 1.35 (1.10, 1.70) | 1.41 (0.389) | 1.30 (1.20, 1.50) |
| Total cholesterol, mmol/L | 4.61 (0.757) | 4.80 (3.98, 5.12) | 4.98 (0.796) | 5.00 (4.50, 5.40) |
| Triglycerides, mmol/L | 1.05 (0.43) | 1.00 (0.70, 1.20) | 1.10 (0.42) | 1.00 (0.90, 1.40) |
| Glucose, mmol/L | 5.16 (0.38) | 5.25 (4.90, 5.43) | 5.26 (0.45) | 5.40 (5.00, 5.60) |
| Insulin, pmol/L | 84 (47.8) | 78.5 (45.5, 104) | 73.0 (40.2) | 61.0 (47.0, 98.0) |
| ASAT, U/L | 24.8 (9.14) | 24.0 (19.0, 26.0) | 22.0 (5.96) | 20.0 (18.0, 25.0) |
| ALAT, U/L | 24.9 (8.3) | 23.5 (20.0, 27.0) | 25.5 (11.6) | 22.0 (17.0, 31.0) |
| Creatinine, **µ**mol/L | 65.9 (11.6) | 67.0 (55.0, 74.2) | 68.7 (12.2) | 72.0 (60.0, 76.0) |
| Resting metabolic rate, kcal/d | 2016 (288) | 2016 (1814, 2116) | 1944 (297) | 1944 (1728, 2188) |
| RQ | 0.79 (0.04) | 0.78 (0.76, 0.81) | 0.78 (0.03) | 0.79 (0.76, 0.80) |
| ^1^All variables are mean (standard deviation) and median (IQR) except row for males. Abbreviations: ALAT, alanine aminotransferase; ASAT, aspartate aminotransferase; BMI, body mass index; LDL, low-density lipoprotein; HDL, high-density lipoprotein; FGF-21, fibroblast growth factor 21; IGF-1, insulin-like growth factor 1; RMR, resting metabolic rate expenditure; RQ, respiratory quotient | | | | |

| Supplemental Table S7. Effects of SAAR on all parameters in the per-protocol population^1^. | | | | |
| --- | --- | --- | --- | --- |
|  | **4 weeks** | | **8 weeks** | |
|  | **β (95 % CI)**  **SAAR vs. control** | **p-value** | **β (95 % CI)**  **SAAR vs. control** | **p-value** |
| Body weight, kg | -0.85 (-1.79, 0.08) | 0.074 | -1.22 (-2.13, -0.30) | 0.010 |
| BMI, kg/m^2^ | -0.25 (-0.56, 0.06) | 0.12 | -0.33 (-0.64, -0.03) | 0.033 |
| Body fat, % | -0.07 (-0.68, 0.53) | 0.81 | 0.01 (-0.58, 0.60) | 0.97 |
| Total fat, kg | -0.32 (-1.06, 0.42) | 0.39 | -0.41 (-1.13, 0.31) | 0.26 |
| Arm fat, kg | -0.03 (-0.15, 0.10) | 0.65 | -0.03 (-0.15, 0.10) | 0.67 |
| Leg fat, kg | -0.11 (-0.41, 0.19) | 0.47 | -0.16 (-0.45, 0.14) | 0.30 |
| Trunk fat, kg | -0.14 (-0.69, 0.41) | 0.61 | -0.2 (-0.74, 0.34) | 0.46 |
| Android fat, kg | 0.02 (-0.1, 0.13) | 0.79 | -0.05 (-0.17, 0.07) | 0.39 |
| Gynoid fat, kg | -0.04 (-0.18, 0.10) | 0.57 | -0.13 (-0.26, 0.00) | 0.057 |
| Visceral adipose tissue, kg | 0.05 (-0.04, 0.14) | 0.28 | 0.02 (-0.06, 0.11) | 0.61 |
| Subcutaneous adipose tissue, kg | -0.02 (-0.12, 0.08) | 0.65 | -0.07 (-0.17, 0.02) | 0.14 |
| Fat free mass, kg | -0.45 (-0.94, 0.05) | 0.08 | -0.72 (-1.20, -0.23) | 0.012 |
| Resting metabolic rate, kcal/d | 56.32 (-22.8, 135) | 0.16 | 14.7 (-62.1, 91.5) | 0.71 |
| Fat oxidation, g/d | 21.4 (2.97, 39.8) | 0.02 | 13.1 (-4.66, 30.9) | 0.15 |
| Carbohydrate oxidation, g/d | -24.8 (-61.6, 12.0) | 0.18 | -18.2 (-53.7, 17.3) | 0.31 |
| RQ | -0.01 (-0.04, 0.01) | 0.16 | -0.01 (-0.03, 0.01) | 0.21 |
| ***Sulfur amino acids*** |  |  |  |  |
| Methionine excretion, mmol/24h | -3.74 (-5.94, -1.54) | 0.001 | -2.69 (-4.89, -0.49) | 0.017 |
| tHcy excretion, mmol/24h | -2.80 (-4.61, -0.99) | 0.003 | -2.93 (-4.74, -1.13) | 0.002 |
| Cystathionine excretion, mmol/24h | -29.4 (-45.7, -13.2) | < 0.001 | -31.8 (-48.1, -15.6) | < 0.001 |
| tCys excretion, mmol/24h | -51.8 (-101, -2.00) | 0.042 | -77.3 (-127, -27.5) | 0.003 |
| Sulfate excretion, mol/24h | -22.3 (-27.1, -17.6) | < 0.001 | -20.2 (-24.9, -15.5) | < 0.001 |
| Plasma methionine, **µmol/L** | -0.41 (-2.09, 1.28) | 0.63 | -1.16 (-2.80, 0.47) | 0.16 |
| Plasma tHcy, **µmol/L** | 1.84 (0.97, 2.71) | < 0.001 | 2.06 (1.21, 2.90) | < 0.001 |
| Plasma cystathionine, **µmol/L** | -0.17 (-0.24, -0.09) | < 0.001 | -0.17 (-0.23, -0.10) | < 0.001 |
| Plasma tCys, **µmol/L** | 12.3 (-3.19, 27.8) | 0.12 | 12.0 (-3.03, 27.1) | 0.12 |
| ***Plasma biomarkers*** |  |  |  |  |
| Triglycerides, mmol/L | -0.01 (-0.20, 0.17) | 0.90 | -0.15 (-0.32, 0.03) | 0.11 |
| Total cholesterol, mmol/L | 0.05 (-0.27, 0.37) | 0.75 | -0.07 (-0.38, 0.24) | 0.66 |
| LDL, mmol/L | -0.01 (-0.27, 0.24) | 0.92 | -0.05 (-0.29, 0.20) | 0.70 |
| HDL, mmol/L | 0.05 (-0.05, 0.15) | 0.34 | 0.05 (-0.04, 0.15) | 0.29 |
| Apolipoprotein B, g/L | 0.01 (-0.07, 0.09) | 0.76 | -0.01 (-0.09, 0.07) | 0.74 |
| Apolipoprotein A1, g/l | 0.03 (-0.06, 0.12) | 0.48 | -0.01 (-0.09, 0.08) | 0.85 |
| Fasting glucose, mmol/L | 0.00 (-0.18, 0.18) | 0.99 | -0.08 (-0.25, 0.10) | 0.39 |
| ***Plasma hormones*** |  |  |  |  |
| Fasting insulin, pmol/L | -1.82 (-19.3, 15.6) | 0.84 | -9.08 (-26.0, 7.87) | 0.29 |
| C-peptide, pmol/L | 5.87 (-91.6, 103) | 0.91 | -58.0 (-152, 36.5) | 0.23 |
| TSH, mU/L | -0.08 (-0.43, 0.26) | 0.64 | -0.30 (-0.64, 0.03) | 0.07 |
| Free T4, pmol/L | 0.12 (-1.17, 1.42) | 0.85 | -0.28 (-1.54, 0.98) | 0.66 |
| FGF-21, pg/mL^2^ | -61.0 (-448, 326) | 0.76 | -60.4 (-439, 318) | 0.75 |
| IGF-1, pg/mL | - | - | -11.3 (-41.4, 18.7) | 0.46 |
| Adiponectin, pg/mL | *-* | *-* | -1.44 (-4.46, 1.58) | 0.34 |
| Gastric inhibitory peptide, pg/mL | -11.0 (-70.0, 48.1) | 0.71 | 24.1 (-33.5, 81.7) | 0.41 |
| Glucagon-like peptide 1, pg/mL | -12.6 (-22.2, -3.03) | 0.010 | -8.20 (-17.6, 1.16) | 0.085 |
| Ghrelin, pg/mL | -7.66 (-49.3, 34.0) | 0.72 | -12.3 (-52.9, 28.3) | 0.55 |
| Leptin, ng/mL | -3.86 (-9.48, 1.75) | 0.18 | -5.92 (-11.4, -0.44) | 0.03 |
| Pancreatic peptide, pg/mL | 41.6 (-26.8, 110) | 0.23 | 48.1 (-18.8, 115) | 0.16 |
| Peptide YY, pg/mL | -6.19 (-19.5, 7.10) | 0.36 | -0.76 (-13.7, 12.2) | 0.91 |
| **Ketone bodies** |  |  |  |  |
| β-hydroxybutyrate, **µmol/L^2^** | 49.2 (-56.3, 154) | 0.36 | 116 (14.4, 219) | 0.026 |
| Acetoacetate, **µmol/L^2^** | 12.02 (-15.1, 39.2) | 0.38 | 27.0 (0.68, 53.3) | 0.044 |
| Acetone, **µmol/L** | 3.14 (-10.6, 16.9) | 0.42 | 3.14 (-10.2, 16.5) | 0.089 |
| Acetate, **µmol/L** | 4.14 (-5.95, 14.2) | 0.65 | 8.47 (-1.32, 18.3) | 0.64 |
| ***Safety markers*** |  |  |  |  |
| ASAT, U/L | 3.98 (-0.20, 8.16) | 0.062 | 0.91 (-3.10, 4.92) | 0.65 |
| ALAT, U/L | 2.41 (-3.98, 8.81) | 0.46 | -1.59 (-7.80, 4.61) | 0.61 |
| Urea, mmol/L | -0.26 (-0.64, 0.11) | 0.16 | -0.10 (-0.46, 0.27) | 0.60 |
| Creatinine, **µmol/L** | 3.83 (1.02, 6.64) | 0.008 | 3.68 (0.94, 6.42) | 0.009 |
| Vitamin B12, pmol/L | 55.6 (-3.71, 115) | 0.066 | 9.88 (-47.8, 67.6) | 0.74 |
| MMA, **µmol/L** | 0.01 (-0.03, 0.04) | 0.76 | 0.03 (-0.01, 0.07) | 0.17 |

| Supplemental Table S8. Estimated marginal mean (95 % CI) body composition in the SAAR and control groups in the per protocol population^1^. | | | | | | |
| --- | --- | --- | --- | --- | --- | --- |
|  | **Group** | **Baseline** | **4 weeks** | **8 weeks** | **p_int 4 weeks_** | **p_int_ _8 weeks_** |
| ***Body weight and composition*** |  |  |  |  |  |  |
| Body weight, kg | Control | 94.5  (91.7, 97.2) | 91.1  (88.3, 93.9) | 89.1  (86.3, 91.9) | 0.074 | 0.010 |
|  | SAAR | 94.5  (91.7, 97.2) | 90.3  (87.5, 93.1) | 87.9  (85.1, 90.7) |  |  |
| BMI, kg/m^2^ | Control | 30.9  (30.2, 31.6) | 29.8  (29.1, 30.5) | 29.1  (28.4, 29.8) | 0.12 | 0.033 |
|  | SAAR | 30.9  (30.2, 31.6) | 29.5  (28.8, 30.2) | 28.7  (28, 29.4) |  |  |
| Body fat, % | Control | 39.6  (38.2, 40.9) | 38.9  (37.5, 40.3) | 38.0  (36.6, 39.4) | 0.81 | 0.97 |
|  | SAAR | 39.6  (38.2, 40.9) | 38.9  (37.5, 40.2) | 38.0  (36.7, 39.4) |  |  |
| Total fat, kg | Control | 35.8  (34.0, 37.7) | 33.9  (32.0, 35.8) | 32.5  (30.6, 34.3) | 0.39 | 0.26 |
|  | SAAR | 35.8  (34.0, 37.7) | 33.6  (31.7, 35.5) | 32.0  (30.2, 33.9) |  |  |
| Arm fat, kg | Control | 3.76  (3.54, 3.99) | 3.68  (3.45, 3.92) | 3.53  (3.30, 3.77) | 0.65 | 0.67 |
|  | SAAR | 3.76  (3.54, 3.99) | 3.66  (3.42, 3.89) | 3.51  (3.27, 3.74) |  |  |
| Leg fat, kg | Control | 11.7  (11.0, 12.4) | 11.3  (10.5, 12) | 10.8  (10.0, 11.5) | 0.47 | 0.30 |
|  | SAAR | 11.7  (11.0, 12.4) | 11.1  (10.4, 11.9) | 10.6  (9.89, 11.3) |  |  |
| Trunk fat, kg | Control | 19.4  (18.1, 20.8) | 18.0  (16.6, 19.4) | 17.2  (15.8, 18.6) | 0.61 | 0.46 |
|  | SAAR | 19.4  (18.1, 20.8) | 17.8  (16.4, 19.2) | 17  (15.6, 18.4) |  |  |
| Android fat, kg | Control | 3.36  (3.08, 3.64) | 3.06  (2.77, 3.34) | 2.92  (2.63, 3.2) | 0.80 | 0.39 |
|  | SAAR | 3.36  (3.08, 3.64) | 3.07  (2.79, 3.36) | 2.87  (2.58, 3.15) |  |  |
| Gynoid fat, kg | Control | 6.13  (5.80, 6.46) | 5.77  (5.44, 6.11) | 5.53  (5.20, 5.87) | 0.57 | 0.06 |
|  | SAAR | 6.13  (5.80, 6.46) | 5.74  (5.40, 6.07) | 5.41  (5.07, 5.74) |  |  |
| Visceral adipose tissue, kg | Control | 1.20  (1.04, 1.36) | 1.02  (0.85, 1.18) | 0.99  (0.82, 1.15) | 0.28 | 0.61 |
|  | SAAR | 1.20  (1.04, 1.36) | 1.07  (0.90, 1.23) | 1.01  (0.84, 1.17) |  |  |
| Subcutaneous adipose tissue, kg | Control | 2.48  (2.26, 2.7) | 2.3  (2.08, 2.53) | 2.17  (1.95, 2.39) | 0.65 | 0.14 |
|  | SAAR | 2.48  (2.26, 2.7) | 2.28  (2.06, 2.5) | 2.10  (1.87, 2.32) |  |  |
| Fat free mass, kg | Control | 58.1  (56.5, 59.7) | 56.8  (55.1, 58.4) | 56.4  (54.7, 58) | 0.08 | 0.012 |
|  | SAAR | 58.1  (56.5, 59.7) | 56.3  (54.7, 57.9) | 55.7  (54, 57.3) |  |  |
| ***Energy expenditure*** |  |  |  |  |  |  |
| RMR, kcal/day^2^ | Control | 1977  (1912, 2044) | 1911  (1840, 1981) | 1914  (1846, 1981) | 0.16 | 0.71 |
|  | SAAR | 1977  (1912, 2044) | 1958  (1887, 2029) | 1927  (1858, 1996) |  |  |
| Fat oxidation, g/day | Control | 126  (116, 136) | 130  (116, 144) | 127  (114, 141) | 0.02 | 0.15 |
|  | SAAR | 126  (116, 136) | 149  (136, 162) | 139  (126, 152) |  |  |
| Glucose oxidation, g/day | Control | 118  (99.2, 137) | 53.2  (27.5, 78.8) | 67.8  (42.2, 93.4) | 0.18 | 0.31 |
|  | SAAR | 118  (99.2, 137) | 74.3  (46.7, 102) | 79.7  (53.7, 106) |  |  |
| Respiratory quotient | Control | 0.78  (0.77, 0.79) | 0.77  (0.75, 0.78) | 0.77  (0.75, 0.78) | 0.16 | 0.21 |
|  | SAAR | 0.78  (0.77, 0.79) | 0.76  (0.74, 0.77) | 0.76  (0.75, 0.78) |  |  |
| ***Plasma sulfur amino acids, µmol/L*** |  |  |  |  |  |  |
| Methionine | Control | 22.6  (21.4, 23.8) | 22.1  (20.6, 23.6) | 22.6  (21.1, 24.0) | 0.63 | 0.16 |
|  | SAAR | 22.6  (21.4, 23.8) | 21.7  (20.3, 23.1) | 21.4  (20, 22.8) |  |  |
| Total homocysteine | Control | 9.98  (9.14, 10.8) | 8.59  (7.62, 9.56) | 8.75  (7.80, 9.70) | < 0.001 | < 0.001 |
|  | SAAR | 9.98  (9.14, 10.8) | 10.4  (9.49, 11.4) | 10.8  (9.86, 11.7) |  |  |
| Cystathionine | Control | 0.24  (0.20, 0.28) | 0.29  (0.24, 0.35) | 0.31  (0.25, 0.36) | < 0.001 | < 0.001 |
|  | SAAR | 0.24  (0.20, 0.28) | 0.13  (0.08, 0.18) | 0.14  (0.09, 0.20) |  |  |
| Total cysteine | Control | 271  (261, 281) | 260  (246, 273) | 265  (253, 278) | 0.12 | 0.12 |
|  | SAAR | 271  (261, 281) | 272  (259, 284) | 277  (265, 290) |  |  |
| ***Sulfur amino acid output*** |  |  |  |  |  |  |
| Methionine, µmol/24h | Control | 10.7  (9.4, 12.1) | 10.3  (8.46, 12.1) | 10.0  (8.23, 11.9) | 0.001 | 0.017 |
|  | SAAR | 10.7  (9.4, 12.1) | 6.52  (4.8, 8.25) | 7.35  (5.63, 9.08) |  |  |
| Total homocysteine, µmol/24h | Control | 9.14  (8.06, 10.2) | 9.12  (7.64, 10.6) | 8.96  (7.48, 10.4) | 0.003 | 0.002 |
|  | SAAR | 9.14  (8.06, 10.2) | 6.32  (4.92, 7.73) | 6.02  (4.62, 7.43) |  |  |
| Cystathionine, µmol/24h | Control | 40.4  (31.6, 49.1) | 37.5  (25.0, 50.1) | 42.1  (29.5, 54.6) | < 0.001 | <0.001 |
|  | SAAR | 40.4  (31.6, 49.1) | 8.10  (-3.79, 20) | 10.2  (-1.65, 22.1) |  |  |
| Total cysteine, µmol/24h | Control | 262  (234, 291) | 309  (269, 348) | 305  (265, 344) | 0.042 | 0.003 |
|  | SAAR | 262  (234, 291) | 257  (220, 294) | 227  (190, 265) |  |  |
| Sulfate, mol/24 h | Control | 18.4  (15.8, 21.0) | 28.5  (24.8, 32.2) | 29.5  (25.6, 33.4) | < 0.001 | < 0.001 |
|  | SAAR | 18.4  (15.8, 21.0) | 7.52  (3.96, 11.1) | 8.75  (5.18, 12.3) |  |  |
| ***Plasma biomarkers*** |  |  |  |  |  |  |
| Triglycerides, mmol/L | Control | 1.11  (0.98, 1.24) | 1.18  (1.01, 1.35) | 1.19  (1.03, 1.35) | 0.90 | 0.11 |
|  | SAAR | 1.11  (0.98, 1.24) | 1.17  (1.01, 1.33) | 1.04  (0.88, 1.20) |  |  |
| Total cholesterol, mmol/L | Control | 4.81  (4.57, 5.06) | 4.16  (3.86, 4.47) | 4.28  (3.98, 4.57) | 0.75 | 0.66 |
|  | SAAR | 4.81  (4.57, 5.06) | 4.22  (3.93, 4.51) | 4.21  (3.92, 4.5) |  |  |
| LDL, mmol/L | Control | 3.04  (2.83, 3.25) | 2.64  (2.39, 2.89) | 2.65  (2.41, 2.89) | 0.92 | 0.70 |
|  | SAAR | 3.04  (2.83, 3.25) | 2.62  (2.38, 2.86) | 2.60  (2.36, 2.84) |  |  |
| HDL, mmol/L | Control | 1.36  (1.27, 1.46) | 1.17  (1.06, 1.28) | 1.20  (1.09, 1.31) | 0.34 | 0.29 |
|  | SAAR | 1.36  (1.27, 1.46) | 1.22  (1.12, 1.33) | 1.25  (1.14, 1.36) |  |  |
| Apolipoprotein B, g/L | Control | 0.88  (0.82, 0.95) | 0.79  (0.72, 0.87) | 0.81  (0.73, 0.88) | 0.76 | 0.74 |
|  | SAAR | 0.88  (0.82, 0.95) | 0.81  (0.73, 0.88) | 0.79  (0.72, 0.87) |  |  |
| Apolipoprotein A1, g/l | Control | 1.44  (1.37, 1.52) | 1.26  (1.17, 1.35) | 1.29  (1.21, 1.38) | 0.48 | 0.85 |
|  | SAAR | 1.44  (1.37, 1.52) | 1.29  (1.20, 1.37) | 1.29  (1.20, 1.37) |  |  |
| Fasting glucose, mmol/L | Control | 5.25  (5.14, 5.36) | 5.14  (4.99, 5.29) | 5.08  (4.94, 5.22) | 0.99 | 0.39 |
|  | SAAR | 5.25  (5.14, 5.36) | 5.14  (5.00, 5.28) | 5.00  (4.86, 5.15) |  |  |
| ***Plasma hormones*** |  |  |  |  |  |  |
| Fasting insulin, pmol/L | Control | 75.1  (64.5, 85.6) | 57.7  (43.3, 72.1) | 60.9  (47.1, 74.7) | 0.84 | 0.29 |
|  | SAAR | 75.1  (64.5, 85.6) | 55.9  (42.3, 69.5) | 51.8  (38.2, 65.4) |  |  |
| C-peptide, pmol/L | Control | 689  (632, 746) | 604  (525, 684) | 631  (555, 706) | 0.91 | 0.23 |
|  | SAAR | 689  (632, 746) | 610  (536, 685) | 573  (498, 647) |  |  |
| TSH, mU/L | Control | 1.47  (1.25, 1.70) | 1.62  (1.32, 1.92) | 1.78  (1.50, 2.07) | 0.64 | 0.074 |
|  | SAAR | 1.47  (1.25, 1.70) | 1.54  (1.26, 1.82) | 1.48  (1.20, 1.76) |  |  |
| Free T4, pmol/L | Control | 16.4  (15.6, 17.1) | 16.7  (15.7, 17.8) | 16.9  (15.9, 17.9) | 0.85 | 0.66 |
|  | SAAR | 16.4  (15.6, 17.1) | 16.9  (15.9, 17.8) | 16.6  (15.6, 17.6) |  |  |
| FGF-21, pg/mL | Control | 574  (460, 716) | 1100  (845, 1430) | 995  (768, 1290) | 0.90 | 0.63 |
|  | SAAR | 574  (460, 716) | 1120  (868, 1440) | 1060  (822, 1370) |  |  |
| IGF-1, pg/mL | Control | 140  (123, 157) | - | 194  (171, 218) | - | 0.46 |
|  | SAAR | 140  (123, 157) | - | 183  (160, 206) |  |  |
| Adiponectin, pg/mL | Control | 20.3  (18.0, 22.6) | - | 20.5  (17.7, 23.4) | - | 0.34 |
|  | SAAR | 20.3  (18.0, 22.6) | - | 19.1  (16.3, 21.9) |  |  |
| Gastric inhibitory peptide, pg/mL | Control | 182  (117, 247) | 183  (111, 256) | 183  (111, 254) | 0.71 | 0.41 |
|  | SAAR | 182  (117, 247) | 172  (102, 243) | 207  (136, 278) |  |  |
| Glucagon-like peptide 1, pg/mL | Control | 63.4  (56.2, 70.5) | 66.4  (57.5, 75.3) | 63.5  (54.8, 72.2) | 0.010 | 0.085 |
|  | SAAR | 63.4  (56.2, 70.5) | 53.8  (45.3, 62.3) | 55.3  (46.8, 63.8) |  |  |
| Leptin, ng/mL | Control | 43.2  (37.5, 49) | 32.6  (26.1, 39.1) | 31.7  (25.3, 38.1) | 0.18 | 0.03 |
|  | SAAR | 43.2  (37.5, 49) | 28.8  (22.4, 35.1) | 25.8  (19.5, 32.1) |  |  |
| Pancreatic peptide, pg/mL | Control | 169  (125, 213) | 146  (87.4, 204) | 156  (99.3, 212) | 0.23 | 0.16 |
|  | SAAR | 169  (125, 213) | 187  (132, 242) | 204  (149, 259) |  |  |
| Peptide YY, pg/mL | Control | 63.2  (51.5, 75) | 71.4  (57.6, 85.3) | 66.3  (52.8, 79.9) | 0.36 | 0.91 |
|  | SAAR | 63.2  (51.5, 75) | 65.2  (51.9, 78.6) | 65.6  (52.2, 78.9) |  |  |
| Ghrelin, pg/mL | Control | 145  (95, 194) | 202  (147, 256) | 186  (132, 240) | 0.72 | 0.55 |
|  | SAAR | 145  (95, 194) | 194  (141, 247) | 174  (120, 227) |  |  |
| ***Ketone bodies*** |  |  |  |  |  |  |
| β-hydroxybutyrate, µmol/L | Control | 24.8  (17.5, 35.3) | 69.0  (41.4, 115) | 55.2  (33.8, 90.1) | 0.36 | 0.026 |
|  | SAAR | 24.8  (17.5, 35.3) | 94.1  (58.1, 152) | 116  (72.1, 186) |  |  |
| Acetoacetate, µmol/L | Control | 15.9  (12.7, 19.9) | 32.2  (23.1, 45.1) | 29.6  (21.4, 40.8) | 0.38 | 0.044 |
|  | SAAR | 15.9  (12.7, 19.9) | 37.2  (27.1, 50.9) | 46.4  (34.1, 63.3) |  |  |
| Acetone, µmol/L | Control | 17.6  (12.4, 22.8) | 26.3  (18.6, 34) | 24.9  (17.5, 32.3) | 0.65 | 0.64 |
|  | SAAR | 17.6  (12.4, 22.8) | 30.5  (23.2, 37.8) | 33.4  (26.3, 40.5) |  |  |
| Acetate, µmol/L | Control | 44.2  (36.8, 51.5) | 53.9  (43.3, 64.5) | 53.9  (43.7, 64.1) | 0.42 | 0.089 |
|  | SAAR | 44.2  (36.8, 51.5) | 57.0  (46.9, 67.1) | 57.0  (47.1, 66.9) |  |  |
| ***Safety markers*** |  |  |  |  |  |  |
| ASAT, U/L | Control | 24.3  (22.1, 26.5) | 22.4  (19.2, 25.6) | 22.9  (19.9, 25.9) | 0.06 | 0.65 |
|  | SAAR | 24.3  (22.1, 26.5) | 26.4  (23.4, 29.5) | 23.8  (20.8, 26.8) |  |  |
| ALAT, U/L | Control | 27.4  (23.6, 31.1) | 28.3  (23.1, 33.5) | 26.3  (21.3, 31.2) | 0.46 | 0.61 |
|  | SAAR | 27.4  (23.6, 31.1) | 30.8  (25.9, 35.6) | 24.7  (19.8, 29.6) |  |  |
| Urea, mmol/L | Control | 4.41  (4.12, 4.7) | 3.31  (2.95, 3.66) | 3.38  (3.03, 3.72) | 0.16 | 0.60 |
|  | SAAR | 4.41  (4.12, 4.7) | 3.04  (2.7, 3.38) | 3.28  (2.94, 3.62) |  |  |
| Creatinine, µmol/L | Control | 70.1  (67.2, 73) | 67.9  (64.7, 71.2) | 66.4  (63.2, 69.6) | 0.008 | 0.009 |
|  | SAAR | 70.1  (67.2, 73) | 71.8  (68.6, 74.9) | 70.1  (66.9, 73.3) |  |  |
| Vitamin B12, pmol/L | Control | 387  (341, 433) | 370  (314, 427) | 407  (352, 462) | 0.066 | 0.74 |
|  | SAAR | 387  (341, 433) | 426  (372, 480) | 417  (363, 471) |  |  |
| MMA, µmol/L | Control | 0.18  (0.16, 0.20) | 0.15  (0.13, 0.18) | 0.16  (0.13, 0.19) | 0.76 | 0.17 |
|  | SAAR | 0.18  (0.16, 0.20) | 0.16  (0.13, 0.19) | 0.18  (0.15, 0.21) |  |  |
| ^1^ Estimated means and p-values are derived from linear mixed models with body mass as the outcome, and group, visit and their interaction term (group × time) at 4 and 8 weeks as predictors. The models were baseline adjusted. The p-value indicates the difference between groups at each timepoint. Abbreviations: BMI, body mass index; SAAR, sulfur amino acid restriction  ^2^ Additionally adjusted for total fat mass | | | | | | |

### 3. Supplementary figures

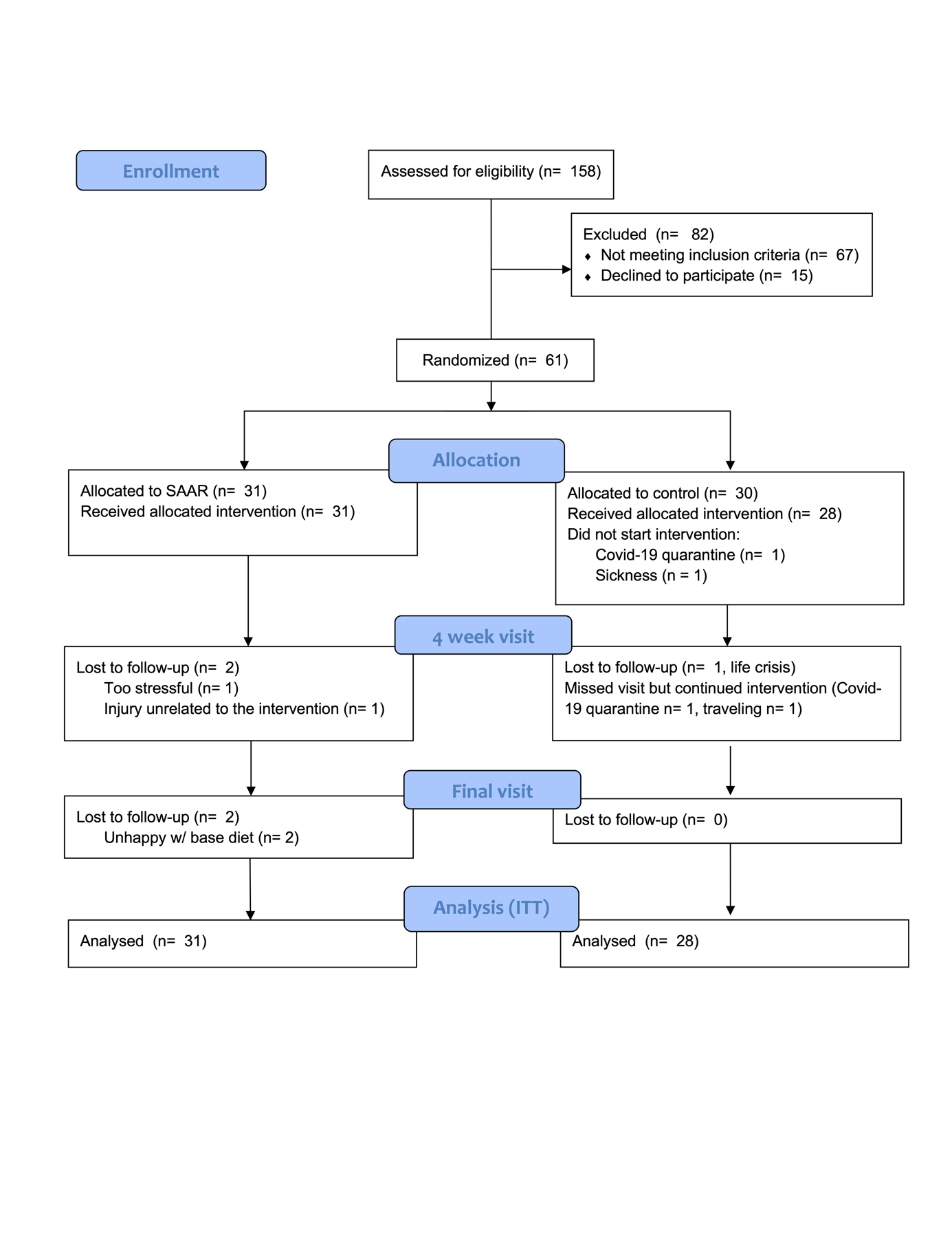

**Supplemental Figure S1.** CONSORT diagram outlining participant flow. Abbreviations: ITT, intention-to-treat; SAAR, sulfur amino acid restriction.
